# Examining the Contribution of Childhood Maltreatment to the Sex Gap in Depression: Insights from the German National Cohort (NAKO)

**DOI:** 10.64898/2026.02.02.26345366

**Authors:** Maja P. Völker, Aina L. Kresken, Jerome C. Foo, Josef Frank, Iris Reinhard, Johanna Klinger-König, Lea Zillich, Diana S. Ferreira de Sá, Rafael Mikolajczyk, Michael Leitzmann, Patricia Fromherz, Lilian Krist, Thomas Keil, Claudia Meinke-Franze, Steffi Riedel-Heller, Halina Greiser, Barbara Bohn, Hermann Brenner, Nadia Obi, Volker Harth, Tobias Pischon, Hans J. Grabe, Klaus Berger, Emanuel Schwarz, Jutta Mata, Stephanie H. Witt, Fabian Streit

## Abstract

**Background:** Childhood maltreatment is a major risk factor for depression and may contribute to sex differences in depression prevalence. We examined sex-specific associations between childhood maltreatment and depression and estimated the proportion of depression cases attributable to maltreatment subtypes.

**Methods:** We analyzed baseline data from 159,045 participants (49.4% female; aged 19–72) in the German National Cohort (NAKO). Childhood maltreatment was assessed via the Childhood Trauma Screener; depression via self-reported physician’s diagnosis and MINI classification (lifetime) and the PHQ-9 (current). Associations, including sex interactions, were modeled using binary logistic regressions. Disparity decomposition analyses and sex-stratified population attributable fractions quantified the contribution of childhood maltreatment to depression.

**Results:** Childhood maltreatment was associated with increased odds of lifetime (OR_physician’s diagnosis_=2.45 [2.38,2.53]; OR_MINI_=2.30 [2.18,2.43]) and current depression (OR=2.90 [2.79,3.02], all p_adj_<0.001). Sex interactions were observed for physician’s diagnosis: physical abuse (p_adj_=0.024) and neglect (p_adj_<0.001) had stronger associations in females (OR_physical abuse_=2.74 [2.59,2.90]; OR_physical neglect_=1.36 [1.28,1.44]) than males (_ORphysical abuse_=2.36 [2.21,2.52]; OR_physical neglect_=1.08 [1.00,1.16]), whereas sexual abuse (p_adj_=0.003) showed stronger associations in males (OR=3.23 [2.91,3.57]) than females (OR=2.61 [2.48,2.75]). Overall, childhood maltreatment accounted for 21.2-26.2% of lifetime and 33.4% of current depression. Population attributable fractions were higher in females than males for lifetime (24.5-28.5% vs. 16.0-20.9%) and current depression (36.1% vs. 28.4%). Emotional subtypes contributed the highest population attributable fractions (up to 10.2%). Childhood maltreatment contributed 19.2-22.4% to the association between sex and depression.

**Conclusion:** Childhood maltreatment accounts for a substantial proportion of depression in both sexes, with stronger overall associations in females. Sex-specific prevention may help reduce depression prevalence.

## Introduction

Childhood maltreatment is a widespread global problem with detrimental consequences for physical and mental health [1]. Commonly, five subtypes are distinguished: sexual, physical, and emotional abuse, and emotional and physical neglect [2]. The estimated global self-reported prevalence rates are 12.7% for sexual abuse, 22.6% for physical abuse, 36.3% for emotional abuse, 18.4% for emotional neglect, and 16.3% for physical neglect [3–5]. The highest childhood maltreatment estimates were reported for South America, while lower estimates were found for Europe and Asia, with the lowest rates in China, the Netherlands and the United Kingdom [6]. A widespread consequence of childhood maltreatment is depression [7,8]. Major depressive disorder (MDD) affects more than 330 million people worldwide and is a leading contributor to the global burden of disease [9,10]. Risk factors also include genetic predisposition, low socioeconomic status, and stressful life events [11]. There is robust evidence linking childhood maltreatment to current and lifetime depression [7,9], especially emotional abuse and neglect [12–14]. A systematic review and meta-analysis found consistent associations of all childhood maltreatment subtypes with depressive disorders, supporting a broad dose-response relationship between childhood maltreatment and affective psychopathology [15]. Individuals who experienced childhood maltreatment are twice as likely to develop depression compared to unaffected individuals. Moreover, affected individuals report earlier onset of depressive symptoms compared to the general population [8,16]. Consistent with these findings, recent studies on childhood maltreatment using data from the German National Cohort (NAKO), a large population-based longitudinal health study of more than 200,000 adults in Germany [17], showed a substantial association of lifetime and current depression with sexual and physical abuse, but an even stronger association with emotional abuse and neglect [18,19].

Sex differences also play a key role in depression: Females are twice as likely to be diagnosed with depression [14,20–22] and report depressive symptoms more frequently [23,24]. There is growing evidence that childhood maltreatment may contribute to this disparity both through differences in the impact of childhood maltreatment and in its prevalence across sexes. Females are affected by childhood maltreatment differently than males in terms of exposure and vulnerability [7,25–29]: Overall, females report childhood maltreatment more frequently [30,31], in particular more severe and non-visible forms such as sexual and emotional abuse [32–35]; while males are more often exposed to physical abuse [36]. Sexual and emotional abuse show stronger associations with internalizing outcomes (e.g., depression) in females compared to males [8], with overall higher susceptibility to psychopathology following early maltreatment exposure [9,21,28]. Additionally, childhood maltreatment is associated with an earlier age of depression onset in females compared to males [8]. The differential exposure is mirrored in the NAKO: 28.7% of females reported a history of childhood maltreatment, compared to 23.5% of males [19]. Specifically, females reported higher rates of sexual (9.7% vs. 2.6%) and emotional (10% vs. 6.2%) abuse, and emotional neglect (8.5% vs. 6.6%) than males, while males reported slightly higher frequencies of physical abuse than females (8.4% vs. 7.8%) [19]. Other German estimates from a nationwide young adult cohort show the same pattern but overall lower prevalence, with 20.9% of females and 16.1% of males reporting childhood maltreatment [37]. Similar to the NAKO, the largest differences emerged for sexual (5.3% vs. 1.7%) and emotional abuse (8.8% vs. 4.8%), while physical abuse (3.9% vs. 3.5%) and physical neglect (8.5% vs. 8.7%) showed minimal sex differences [37].

Although the overall link between childhood maltreatment and depression is well-established, the evidence on how specific subtypes affect depression risk in males and females remains inconsistent and sometimes contradictory [24,30,31]: While most studies find no sex differences [31], MacMillan et al. found a stronger association between physical abuse and depression in females than males [32]. Thomas et al. reported significant positive correlations between emotional abuse and physical neglect with depression for females, but not for males [38]. In a study by Zhou et al. [9,21,28], all subtypes were associated with depression in females, while in males, only emotional abuse and neglect were associated with depression. Clarifying subtype-specific associations is essential for understanding the mechanisms through which childhood maltreatment contributes to depression risk across sexes.

One way to quantify the contribution of childhood maltreatment to depression is Population Attributable Fractions (PAFs; Levin, 1953). PAFs range from 0% to 100% and estimate the share of disease cases in a population that can be attributed to exposure to a specific risk factor [39], corresponding to the potential reduction in the disease proportion if exposure to this risk factor was eliminated [39,40]. PAFs are determined by both the strength of the association between risk factor and outcome, and the prevalence of the risk factor, and can be stratified by sex to investigate sex-specific contributions. Originally developed to assess the impact of a single, independent risk factor on disease burden, PAFs have been expanded to account for multiple, potentially correlated risk factors, thereby better reflecting real-world exposure patterns [41].

Recently, this approach was applied to quantify the contribution of childhood maltreatment and its subtypes to depression: Li et al. [39] estimated that half of MDD cases worldwide could be attributed to childhood maltreatment when the subtypes are considered collectively. However, PAF varied substantially across maltreatment subtypes, with emotional abuse and neglect often showing the largest contributions [39,42]. While these studies did not examine sex-specific differences, other research suggests that these contributions vary between males and females [40,43]. One study in Australia found that a greater proportion of lifetime depression in females (22.8%) than males (15.7%) could be attributed to sexual and physical abuse [44]. Moreover, Xiao et al. [43] investigated all five maltreatment subtypes and sex differences among students in China, but limited their study to current depressive symptoms. Nevertheless, the results showed comparable patterns to those for lifetime depression by Li et al. [39] and Moore et al. [44], with individual contributions of each subtype to depression (6.1%–29.4%) being consistently higher for females. Notably, all of these studies relied on a single definition of depression, leaving it unclear whether subtype-specific contributions differ across multiple depression outcomes. Additionally, most studies only consider specific subtypes, preventing direct comparison of their relative contributions to depression burden in both sexes.

The overall aims of the present study were to assess whether: a) the subtypes of childhood maltreatment differ in their association with lifetime and current depression in males and females and b) sex differences in both the strength of these associations and the prevalence of the childhood maltreatment subtypes result in sex differences in the share of depression that can be attributed to childhood maltreatment (Table 1). We disentangle whether sex differences in depression burden reflect differences in exposure prevalence, effect sizes, or their combined impact. For the present analyses, baseline data of the NAKO were used. To our knowledge, this is the largest study investigating sex-specific associations of childhood maltreatment with depression and the first to provide population-based estimates for Germany.

**Table 1:** Overview of Statistical Analyses for Each Hypothesis with Predictor, Outcome, and sex-specific analysis.

| Aim | Statistical Analysis | Independent Variable | Dependent Variable | Assessment of sex differences |
| --- | --- | --- | --- | --- |
| 1 | Binary logistic regression with interaction term | Childhood maltreatment and the five subtypes [binary: 'yes', 'no'] | Physician's diagnosis of depression<br>[binary: 'physician's diagnosis', 'no physician's diagnosis'] | Interaction variable sex<br>[binary: 'male', 'female'] |
|  |  |  | MINI classification<br>[binary: 'positive MINI classification', 'negative MINI classification'] |  |
| | | | Current depressive symptoms (PHQ-9 cut-off $\geq 10$ [binary: 'current depressive symptoms', 'no current depressive symptoms']) | |
| 2 | Population attributable fractions | Five subtypes of childhood maltreatment [binary: 'yes', 'no'] | Physician's diagnosis of depression<br>[binary: 'physician's diagnosis', 'no physician's diagnosis'] | Stratification by sex [binary: 'male', 'female'] |
|  |  |  | MINI classification<br>[binary: 'positive MINI classification', 'negative MINI classification'] |  |
| | | | Current depressive symptoms (PHQ-9 cut-off $\geq 10$ [binary: 'current depressive symptoms', 'no current depressive symptoms']) | |
| 3 | Decomposition analysis | Independent variable: sex [binary: 'male', 'female']<br>Mediators: five maltreatment subtypes [binary: 'yes', 'no'] | Physician's diagnosis of depression<br>[binary: 'physician's diagnosis', 'no physician's diagnosis'] | Assessed via mediation model (differential exposure and mediated interaction)<br>[binary: 'male', 'female'] |
|  |  |  | MINI classification<br>[binary: 'positive MINI classification', 'negative MINI classification'] |  |
| | | | Current depressive symptoms (PHQ-9 cut-off $\geq 10$ [binary: 'current depressive symptoms', 'no current depressive symptoms']) | |

*Note.* PHQ-9 = Patient Health Questionnaire 9. MINI = Mini-International Neuropsychiatric Interview. Round brackets indicate the coding of a variable. Square brackets specify the measurement scale and levels of the variable. Regressions were calculated with interactions for the whole sample and sex-stratified without the

## Methods

The present analysis used data from the baseline assessment of the NAKO, a prospective, population-based, long-term cohort study [17] that investigates the causes, risk factors, and mechanisms of common diseases.

### Sample

Between 2014 and 2019, a total of 205,415 individuals were recruited from 18 research centers across Germany, randomly selected from population registries based on age and sex [17,45]. Administrative sex was assessed from registry data at the timepoint of recruitment. The recruited sample comprises participants aged 19 to 72 years, with a higher representation of individuals in the age groups ≥40 years. All participants provided written informed consent before participating in the study. Participants were excluded if they were unable to provide explicit consent, answer questions due to insufficient knowledge of German, or did not complete a minimum set of questionnaires and assessments, resulting in a final sample of 205,053 individuals [17]. All participants underwent a Level-1 (L1) program including interviews and touchscreen questionnaires, physical and medical examinations, and the collection of biomaterial [17]. A subset of ∼28% participants underwent a more in-depth Level-2 (L2) assessment. The study was approved by the local ethics committees of all involved study centers and conducted in accordance with the Declaration of Helsinki.

### Measures

#### Self-Reported Physician’s Diagnosis of Depression

For lifetime depression, in a standardized face-to-face interview, participants were asked whether they had ever received a diagnosis of depression from a physician or psychotherapist (from now on: physician’s diagnosis) [23].

#### MINI Depression Classification

Additionally, for lifetime depression, the Major Depression Module of the Mini-International Neuropsychiatric Interview (MINI, German v5.0.0) was used [46]. All participants received a filter question (‘Have you had periods of two weeks or more during your life when you felt depressed or disinterested?’). If affirmed, they were asked about the two cardinal symptoms of depression (i.e., depressed mood, loss of interest or pleasure). If any cardinal symptom was affirmed, only L2 participants received the remaining MINI questions about symptomatology and impairment. For present MINI analyses, only L2 participants with a complete MINI classification were used and dichotomized (positive/negative MINI classification).

#### Patient Health Questionnaire

The depression module of the Patient Health Questionnaire 9 (PHQ-9) [47], comprising nine questions, was used to measure self-reported depressive symptoms over the past two weeks. A sum score ranging from 0 to 27 was calculated, reflecting the severity of current depressive symptoms [23]. The widely used PHQ-9 cut-off score of ≥ 10 (i.e., moderate to severe depressive episode) was used for all analyses [48].

#### Childhood Trauma Screener

The German version of the Childhood Trauma Screener (CTS) [49], an ultra-short version of the Childhood Trauma Questionnaire (CTQ) [2], was used to determine the presence of the five childhood maltreatment subtypes using a single item per subtype [49]. Responses to the questions were provided on a five-point Likert scale [50]. If subjects reported any subtype of childhood maltreatment, it was scored as *any maltreatment*. For the main analyses, *any maltreatment* and all subtypes were analyzed as binary variables (‘no/low trauma’ or ‘moderate/severe trauma’) based on established thresholds [51] (see Supplementary Table S1). In an additional sensitivity analysis, the subtypes were analyzed as five-level categorical variables: 1 = never, 2 = rarely, 3 = sometimes, 4 = often, 5 = very often.

#### Education

Participants’ education levels were categorized according to the International Standard Classification of Education 97 (ISCED97) [52]. Subjects were grouped into 3 categories: low (level 1/2), intermediate (level 3/4), and high (level 5/6) education [53]. An additional category, ‘in progress’ (level 0), was included for participants, primarily under 30 years of age, who were still engaged in education or training at the time of the study.

#### Relative Income Position

The relative income position assesses the income position of the individual relative to the overall median income in Germany. It was derived from the monthly net income and financial need of the household based on the equivalized disposable income of the European Union Statistics on Income and Living Conditions [54]. The relative income position is categorized into five levels from 1: <60% (at risk of poverty) to 5: 150% and higher as provided by the socio-economic NAKO expert group [55].

## Statistical Analyses

Statistical analyses were conducted using R software (v4.4.0). All analyses were adjusted for the following sociodemographic variables: age in years, age² (to account for both linear and non-linear age effects), and education level [18], with sex used solely as a stratification/interaction variable (see below). Education level and relative income position were included as demographic covariates to facilitate comparison with previous work [18], while recognizing that both may be downstream consequences of childhood maltreatment and should therefore not be interpreted as classical confounders in the estimated associations [56,57]. Only subjects with complete information on age (n = 204,725), sex (n = 204,725), education level (n = 190,290), physician’s diagnosis of depression (n = 203,152), PHQ-9 (n = 189,340), and CTS (n = 172,038); and for the L2 subset analyses, with a complete MINI (n = 57,803), were included in the analyses (see Figure 1).

**Figure 1.**
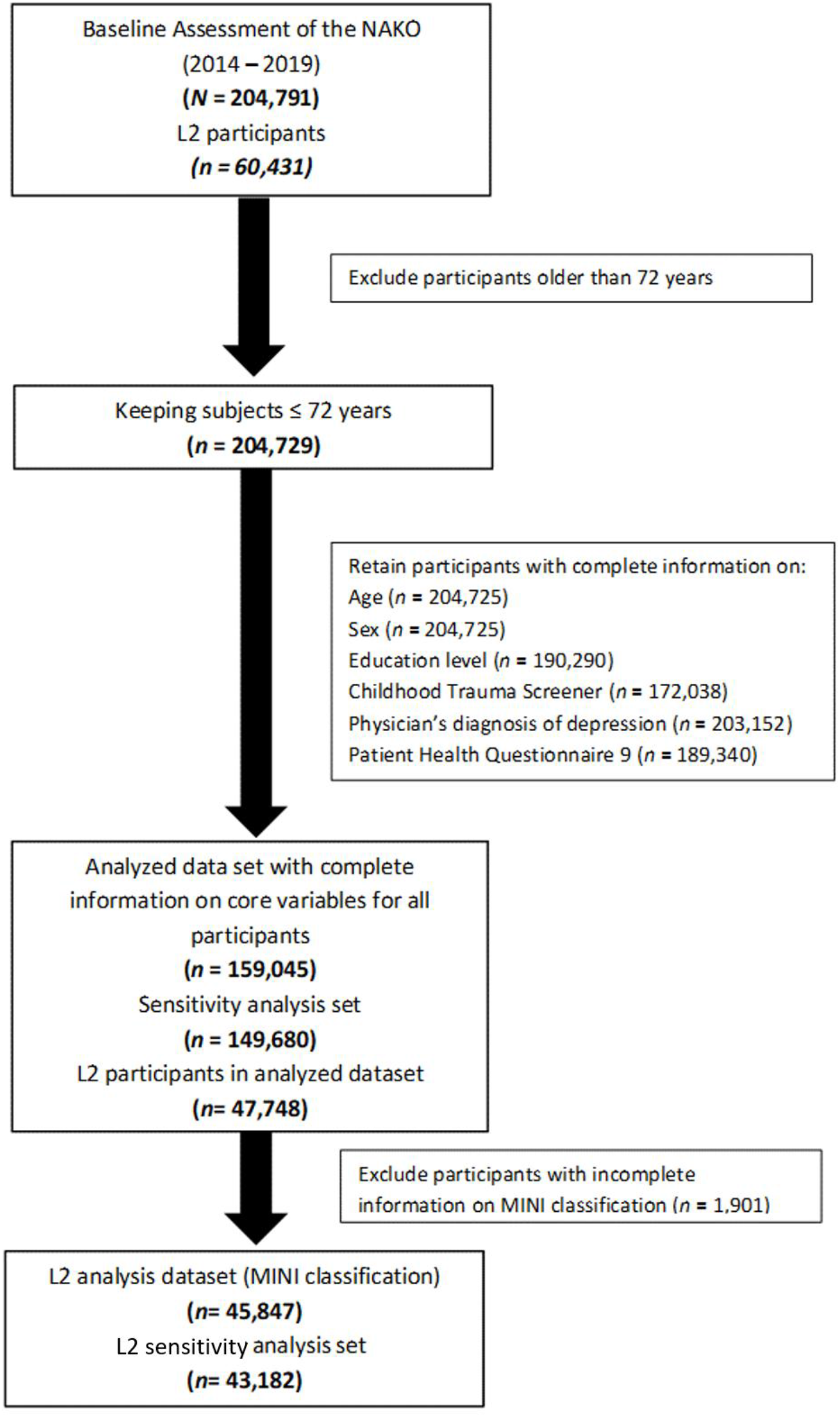
Flow Chart of Sample Selection Displaying Exclusion Criteria *Note:* The figure illustrates the number of excluded subjects and the resulting sample sizes due to missing values. NAKO = German National Cohort. MINI = Mini-International Neuropsychiatric Interview

To assess descriptive sex differences across all included variables, independent samples t-tests (for continuous variables) and ?²-tests (for categorical variables) were used to compare group means and distributions. Corresponding effect sizes were calculated using odds ratios (ORs) for categorical variables, Cohen’s *d [58]* for continuous variables, and Cramer’s *V* [59] for ordinal variables (see Table 2).

**Table 2:** Descriptive Statistics and Association Estimate of All Used Variables for the Total Sample and Separated by Sex.

|  | Total<br>(*N* = 159,045) | Males<br>(*n* = 80,519) | Females<br>(*n* = 78,526) | Association estimate<br>for sex difference | *p*-Value |
| --- | --- | --- | --- | --- | --- |
| Age (mean ± SD) | 48.6 (12.8) | 48.9 (12.9) | 48.3 (12.8) | *d* = 0.05<sup>a</sup> | < 0.001** |
| Sex (male) | 50.6% | - | - | - | - |
| Education | - | - | - | *V* = 0.10<sup>b</sup> | < 0.001** |
| Education high | 56.6% | 61.6% | 51.6% | - | - |
| Education medium | 39.2% | 34.7% | 43.8% | - | - |
| Education low | 1.7% | 1.3% | 2.1% | - | - |
| Education in progress | 2.5% | 2.5% | 2.6% | - | - |
| Physician's diagnosis of depression | 13.9% | 10.1% | 17.7% | *OR* = 1.91 [1.85, 1.96] | < 0.001** |
| MINI classification<sub>1</sub> | 14.6% | 11.9% | 17.6% | *OR* = 1.58 [1.50,1.67] | < 0.001** |
| PHQ-9 ≥ 10 | 7.5% | 5.9% | 9.0% | *OR* = 1.58 [1.52,1.64] | < 0.001** |
| CTS Any Maltreatment | 25.7% | 23.2% | 28.3% | *OR* = 1.31 [1.28,1.34] | < 0.001** |
| CTS Sexual Abuse | 6.0% | 2.5% | 9.5% | *OR* = 4.08 [3.88,4.29] | < 0.001** |
| CTS Physical Abuse | 8.0% | 8.3% | 7.7% | *OR* = 0.93 [0.89,0.96] | < 0.001** |
| CTS Emotional Abuse | 8.0% | 6.2% | 10.0% | *OR* = 1.68 [1.62,1.74] | < 0.001** |
| CTS Emotional Neglect | 7.4% | 6.4% | 8.3% | *OR* = 1.32 [1.27,1.37] | < 0.001** |
| CTS Physical Neglect | 9.7% | 9.8% | 9.7% | *OR* = 0.99 [0.96,1.03] | 0.69 |

*Note.* MINI = Mini-International Neuropsychiatric Interview. PHQ-9 = Patient Health Questionnaire. CTS = Childhood Trauma Screener. *OR* = Odds Ratio. *N*, total number of participants in the sample; *n*, number of participants in sex-stratified groups. Descriptive results are given as mean values (standard deviation) for continuous variables. For categorical variables (physician's diagnosis, MINI, binary PHQ-9, and binary CTS), the values display frequencies (% yes). T-tests (continuous) and $\chi^2$ -tests (categorical) for independent samples were performed to compare means and frequencies between the sexes. *OR*s with 95% confidence intervals for categorical variables, Cohen's *d* for continuous variables, and Cramér's *V* for ordinal variables are reported as effect sizes. \*\* *p* < .001.
<sub>1</sub>Data for MINI classification was available for 45,847 participants.
<sup>a</sup>very small effect. <sup>b</sup>small effect.

To assess whether sex moderates the association between any childhood maltreatment or its subtypes and depression, binary logistic regressions were conducted with childhood maltreatment (*any maltreatment* and all subtypes) as predictors and binary depression measures (physician’s diagnosis, MINI classification, PHQ-9) as outcomes with sex-by-maltreatment interaction terms (resulting in 18 regression models: 6 childhood maltreatment variables × 3 depression outcomes). Additionally, 36 sex-stratified models (6 childhood maltreatment variables × 3 outcomes × 2 sexes) were calculated separately to examine the simple main effects of each childhood maltreatment variable on depression within each sex. A Bonferroni correction across 36 tests was applied. All interaction and sex-stratified models were adjusted for age, age², and education. Sensitivity analyses were conducted with relative income position as additional covariate to further account for socioeconomic status. As additional sensitivity analysis, models were calculated for physician’s diagnosis and PHQ-9 as outcomes in the L2 subset to assess whether the results differed from those obtained in the full sample; and regression models were calculated with CTS subtypes as five-level predictors to account for potential dose-response effects. Bonferroni corrections were applied analogous to the primary analyses (across 18 tests for interaction models, across 36 tests for sex-stratified models).

PAFs [60] were computed to estimate the proportion of depression cases in the cohort that could be attributable to childhood maltreatment. Levin’s formula (1953) was applied,

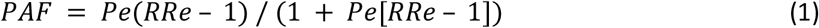

where P_e_ is the prevalence of the risk factor in the population and RR_e_ is the risk ratio of exposure to the risk factor [61]. To demonstrate that the two methods produce equivalent PAF estimates in our study setting, PAFs were also calculated with Miettinen’s formula,

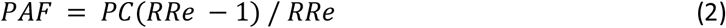

where P_c_ is the proportion of cases exposed to the risk factor in the population and RR_e_ is the risk ratio of exposure to the risk factor. To calculate the PAFs, 15 independent logistic regression models were calculated—one for each combination of the five maltreatment subtypes and the three depression outcomes (physician’s diagnosis, MINI Classification, PHQ-9). Each model was adjusted for age, age², and education, and was run separately for the complete sample and stratified by sex. Sensitivity analyses were performed with additional adjustment for relative income position as a proxy for socioeconomic status; and models were estimated using physician’s diagnosis and PHQ-9 as outcomes in the L2 subset to assess whether the results differed from those obtained in the full sample. Estimated marginal probabilities were obtained from those regression analyses using the R package emmeans [62]. Based on these estimated marginal probabilities, risk ratios (RRs) were calculated. These RRs were used to compute the PAFs [63]. Subsequently, individual PAFs were calculated for each subtype, for the total sample, and stratified by sex. To account for the intercorrelation of maltreatment subtypes, individual PAFs were weighted to avoid overestimating their contribution to the overall PAF [61]. This weighting approach was based on tetrachoric correlations and principal component analysis (PCA) communalities as a pragmatic method to account for overlap among correlated binary risk factors and to reduce double counting of shared attributable burden; however, it is not uniquely determined and does not represent a causal decomposition. The weighting component was determined based on the communalities of the subtypes, i.e., the shared proportion of variance among subtypes. These were derived by computing tetrachoric correlations between the binary childhood maltreatment variables, followed by a PCA. Only the first component had an eigenvalue greater than 1, indicating a reasonable explanatory power; therefore, only its loadings were used to calculate the weights. Then the sum of squares of all factor loadings was calculated to extract the communalities. The weighting factor was determined according to its communality (weight (w) = 1 – communality). Afterwards, the individual PAFs were multiplied by their respective weights and aggregated into a comprehensive overall PAF

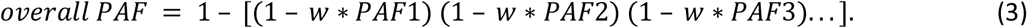

Finally, individual weighted PAFs were recalculated by multiplying them by the overall PAF [61]. All applied statistical analyses are listed in Table 1. To evaluate the significance of sex differences in the sex-stratified PAF estimates (18 comparisons: overall PAF, and five subtype PAFs for physician’s diagnosis, MINI classification and current depression), the difference between males and females was calculated for each of 5,000 bootstrap resamples (i.e., *PAF_males_* – *PAF_females_*). Then, *p*-values were calculated based on the empirical bootstrap distribution of this difference. Specifically, for each of 5,000 bootstrap resamples, the difference between conditions was recalculated, yielding a distribution of bootstrap differences. The p-value was defined as the proportion of bootstrap samples in which the estimated difference had the opposite sign to the observed original difference. To obtain a two-sided p-value, this proportion was multiplied by two. In cases where no bootstrap estimates in the opposite direction to the original observations were obtained, p-values corresponded to *p* < 0.0004 (i.e., fewer than 2/5,000). Frequencies, means, ORs, marginal risks, PAFs, and RRs are reported with unadjusted 95% confidence intervals (CI). For the PAFs, 95% CIs were obtained using bootstrapping with 5,000 resamples separately for each sex (2.5% and 97.5% quantiles) [64]. As a sensitivity analysis, we estimated the total PAF across all childhood maltreatment subtypes using a joint-exposure approach. We defined a single categorical exposure variable representing all possible combinations of the five childhood maltreatment subtypes, such that each participant was assigned to one mutually exclusive exposure profile. This parameterization avoids overlap and potential double counting across correlated risk factors. We then fitted a regression model including this joint exposure variable and relevant covariates, and estimated the total joint PAF by comparing the mean predicted outcome risk under the observed exposure distribution with the mean predicted risk under a counterfactual scenario in which all participants were assigned to the jointly unexposed profile. Formally, the total joint PAF was calculated as

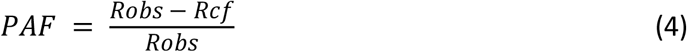

where R_obs_ is the observed mean predicted risk and R_cf_ is the mean predicted risk under the jointly unexposed counterfactual. This analysis was intended to evaluate the robustness of the overall attributable burden to the method used to combine correlated exposures, rather than to distribute burden across individual risk factors.

Bonferroni correction was applied to the primary analyses to adjust p-values for 18 comparisons (i.e., three depression measures × six childhood maltreatment measures: *any maltreatment* and five childhood maltreatment subtypes), using the number of tested sex interactions and sex differences in PAFs, respectively [65]. Bootstrapped *p*-values for the PAF comparisons, < 0.0004, are reported as the corresponding adjusted *p*-value: *p_adj_* < 0.0072. Effects in the sex-stratified regression analyses were corrected for 36 comparisons using the Bonferroni method (i.e., three depression outcomes × six maltreatment measures × two sexes.), and adjusted p-values (*p_adj_*) were reported. Sensitivity analyses and PAF calculations with Miettinen’s formula were conducted to assess robustness and equivalence of estimates and were therefore not treated as independent tests requiring further multiple testing correction. Graphics were created with the R package *ggplot2 [66]*.

To assess the extent to which sex differences in childhood maltreatment prevalence or sex-specific associations between childhood maltreatment and depression account for the higher prevalence of depression in females, separate decomposition models were estimated for each depression outcome. Analyses were based on logistic regression models including sex, the five childhood maltreatment subtypes, age, age², education, and interactions between sex and each maltreatment subtype. Childhood maltreatment subtypes were analyzed simultaneously, thereby accounting for their intercorrelations. The decomposition was performed using standardized predicted probabilities derived from the fitted models. First, the female–male difference in predicted depression risk was estimated under the observed data. Counterfactual predictions were then generated by replacing the observed joint distribution of childhood maltreatment among females with the corresponding distribution observed among males, while leaving all other covariates unchanged, to estimate the contribution of differential exposure. Then, sex-by-maltreatment interaction terms were removed from the prediction model, thereby constraining the associations between childhood maltreatment and depression to be identical for females and males, to estimate the contribution of sex-specific associations. Differential exposure and effect-modification components were calculated using a Shapley decomposition, averaging estimates across the two possible sequences of standardizing the childhood maltreatment distribution and removing interaction effects. Results are presented as absolute risk differences and as the proportion of the observed female–male difference in depression attributable to differential exposure, effect modification, and the remaining unexplained component.

This analysis was preregistered on the Open Science Framework (OSF; https://osf.io/ezjyv) [67]. Bootstrapped analyses estimating confidence intervals of the PAFs, *p*-values for PAF differences, and the additional calculation of PAFs with Miettinen’s formula, sensitivity and decomposition analyses were added to the analysis plan after preregistration. The number of comparisons used for Bonferroni correction of the sex-stratified main effects was revised from the preregistered 18 tests to 36 tests to better capture the actual number of tests.

## Results

### Descriptive Statistics

Descriptive statistics are presented in Table 2. The final sample included 159,045 participants (mean age = 48.6 years, SD = 12.8; 49.4% female; see Figure 1, and Supplementary Table S2 for valid cases and missings of dependent and independent variables). Overall, 13.9% (17.7% of females, 10.1% of males) of the sample reported a physician’s diagnosis and 7.5% (9.0% of females, 5.9% of males) reported current depressive symptoms. Of the L2 participants, 14.6% (17.6% of females, 11.9% of males) received a MINI classification. Childhood maltreatment was more common among females (28.3%) than males (23.2%). The prevalence of maltreatment subtypes across both sexes ranged from 6.0% to 9.7%. Females reported considerably higher frequencies of sexual abuse, emotional abuse, and emotional neglect. Conversely, males were more often exposed to physical abuse than females. For all variables of interest, significant sex differences were observed for group means and distributions (all *p* < 0.001) except for physical neglect (*p* = 0.69; see Table 2).

### Associations of Childhood Maltreatment with Depression and Potential Sex Interactions

In the complete sample, *any maltreatment* showed a significant association with lifetime depression (OR_physician’s diagnosis_ = 2.45, 95% CI [2.38, 2.53]; OR_MINI_ = 2.30, 95% CI [2.18, 2.43], all p_adj_ < 0.001) and current depressive symptoms (OR = 2.90, 95% CI [2.79, 3.02], p_adj_ < 0.001). A significant sex interaction was observed for physician’s diagnosis (β = 0.13, SE = 0.03, z = 4.23, p_adj_ < 0.001) indicating a stronger association between *any maltreatment* and physician’s diagnosis in females (OR = 2.53, 95% CI [2.43, 2.62]), compared to males (OR= 2.19, 95% CI [2.09, 2.30]), but no sex x *any maltreatment* interaction was found for MINI classification (β = 0.10, SE = 0.06, z = 1.79, p_adj_ = 1.000; OR_females_ = 2.36, 95% CI [2.19, 2.54]; OR_males_ = 2.13, 95% CI [1.95, 2.31]), and current depressive symptoms (β = 0.04, SE = 0.04, z = 0.90, p_adj_ = 1.000; OR_females_ = 2.88, 95% CI [2.73, 3.03]; OR_males_ = 2.80, 95% CI [2.63, 2.97]; Figure 2, Supplementary Tables S3-S6). Marginal risks are shown in Supplementary Figures S1-S3. Sensitivity analyses with SES and in the L2 subset are consistent with these results (Supplementary Tables S7-S14).

**Figure 2.**
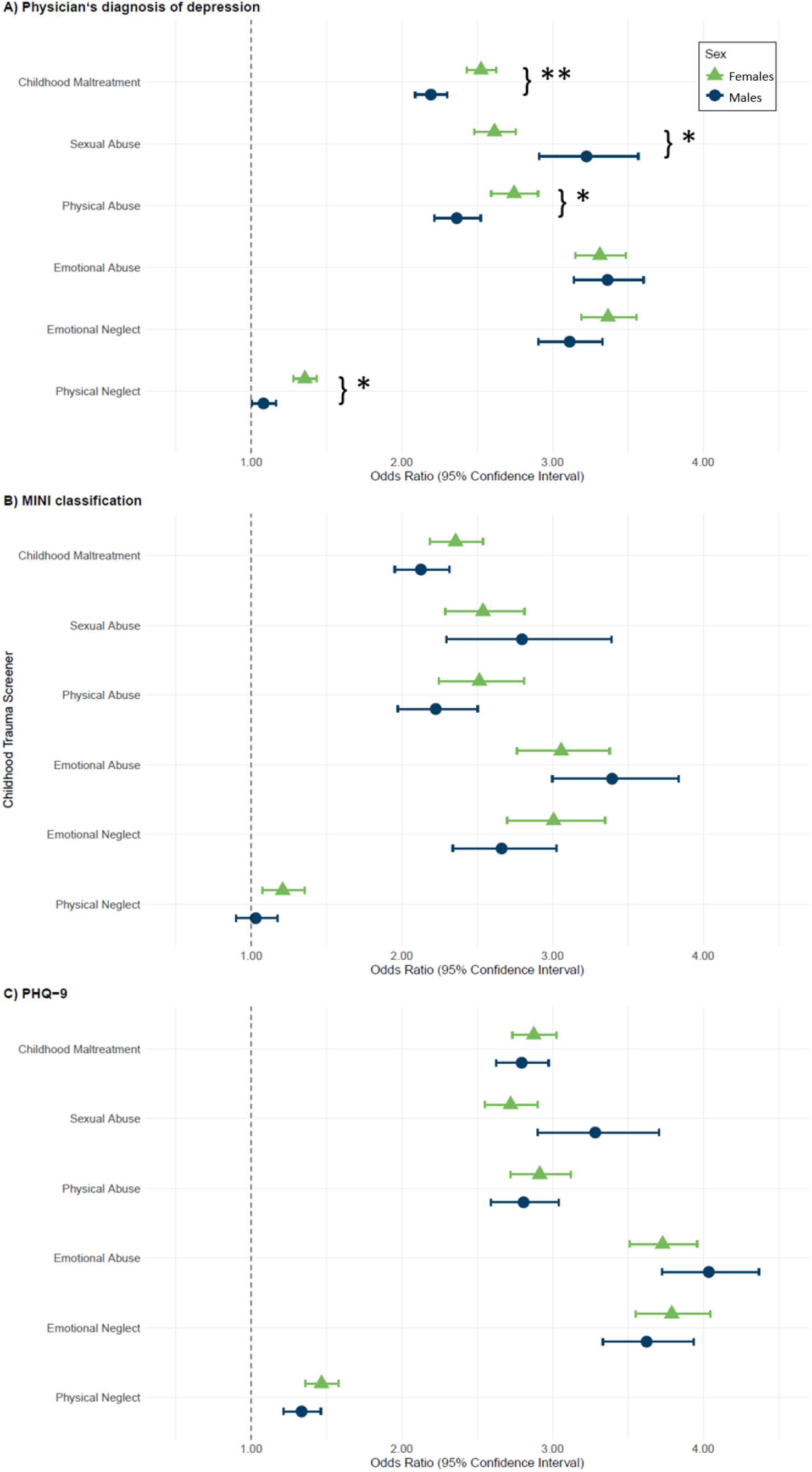
Odds Ratios of Childhood Maltreatment and its Subtypes with Lifetime Depression and Current Depressive Symptoms, Stratified by Sex *Note:* Odds ratios are presented with 95% confidence intervals. ** p_adj_ < 0.001. * p_adj_ < 0.05 for the interaction effect

### Associations of Childhood Maltreatment Subtypes with Depression and Potential Sex Interactions

Each maltreatment subtype was significantly, but with varying association strengths, associated with an increased frequency of lifetime and current depression across sexes (all p_adj_ < 0.001) except for the association between physical neglect and lifetime depression in males (physician’s diagnosis: p_adj_ = 1.000, MINI: p_adj_ = 1.000).

Significant sex interactions in physician’s diagnosis were observed for sexual abuse (β = –.22, SE = 0.06, z = –3.78, p_adj_ = 0.003) with stronger associations in males (OR = 3.23, 95% CI [2.91, 3.57]) than in females (OR = 2.61, 95% CI [2.48, 2.75]), and for physical abuse (β = 0.14, SE = 0.04, z = 3.21, p_adj_ = 0.024), and physical neglect (β = 0.21, SE = 0.05, z = 4.41, p_adj_ < 0.001) with stronger associations in females (OR_physical abuse_ = 2.74, 95% CI [2.59, 2.90], OR_physical neglect_ = 1.36, 95% CI [1.28, 1.44]) than males (OR_physical abuse_ = 2.36, 95% CI [2.21, 2.52], OR_physical neglect_ = 1.08, 95% CI [1.00, 1.16]). All interactions for MINI classification, and current depression were consistent in direction with those for physician’s diagnosis; however, they did not reach significance (all p_adj_ > 0.05; see Figure 2, Supplementary Tables S3-S6). Marginal risks are shown in Supplementary Figures S1-S3. Sensitivity analyses with SES are consistent with these results (Supplementary Tables S7-S10). The associations observed in the sensitivity analyses with the L2 subset are consistent (Supplementary Tables S11-S14); however, the association between physical neglect and physician’s diagnosis was not significant (Supplementary Table S11). Sensitivity analyses using the five-level CTS subtypes largely confirmed the main findings. Across males and females, higher frequencies of childhood trauma were generally associated with progressively greater odds of depression, indicating clear dose–response relationships, particularly for emotional abuse and emotional neglect (Supplementary Tables S15-S16). In addition, the interaction models revealed some level-specific associations that were not apparent in the binary models (e.g., emotional neglect showed significant sex interactions for physician’s diagnosis and PHQ-9), suggesting that dichotomization may have obscured heterogeneity across exposure levels (Supplementary Table S17).

### Population Attributable Fractions for Depression Overall and Stratified by Sex

The overall PAF for the share of lifetime depression associated with the five maltreatment subtypes across the whole sample was 26.2% for physician’s diagnosis (95% CI [25.40, 27.08]) and 21.2% for MINI classification (95% CI [19.76, 22.71]). The estimates for individual subtypes, weighted by their communalities, ranged from 1.5% to 7.5% for physician’s diagnosis and from 0.8% to 6.4% for MINI classification, with the highest PAFs for emotional abuse (physician’s diagnosis: 7.5%; MINI: 6.4%) and neglect (physician’s diagnosis: 6.5%, MINI: 5.1%; Supplementary Table S18). Sex-stratified analyses (Figure 3, Supplementary Tables S19-S20) revealed significantly higher overall PAFs in females than males (all p_adj_ < 0.007): Among females, 28.5% (95% CI [27.3%, 29.7%]) for physician’s diagnosis and 24.5% (95% CI [22.4%, 26.6%]) for MINI classification were associated with childhood maltreatment. For males, 20.9% (95% CI [19.6%, 22.3%]) of physician’s diagnosis and 16.0% of MINI classification (95% CI [14.1%, 18.0%]) were linked to childhood maltreatment. In females, individual PAFs for subtypes ranged from 2.0% to 7.9% for physician’s diagnosis and 1.2% to 6.8% for MINI classification, and in males, from 0.6% to 6.2% for physician’s diagnosis and 0.2% to 5.4% for MINI classification. For both sexes, emotional abuse and neglect, and for females also sexual abuse, showed the largest PAF estimates. Emotional (PAF_females_=7.9%; PAF_males_=6.2%) and sexual (PAF_females_= 6.7%; PAF_males_=2.9%) abuse, and physical (PAF_females_=2.0%; PAF_males_=0.6%) and emotional neglect (PAF_females_=6.8%; PAF_males_=6.0%) were associated with significantly higher PAF estimates for physician’s diagnosis in females than in males (all p_adj_ < 0.005). Physical abuse was the only subtype to not show a significant difference for physician’s diagnosis between males and females (PAF_females_=5.1%; PAF_males_=5.2%, p_adj_=1.000).

**Figure 3.**
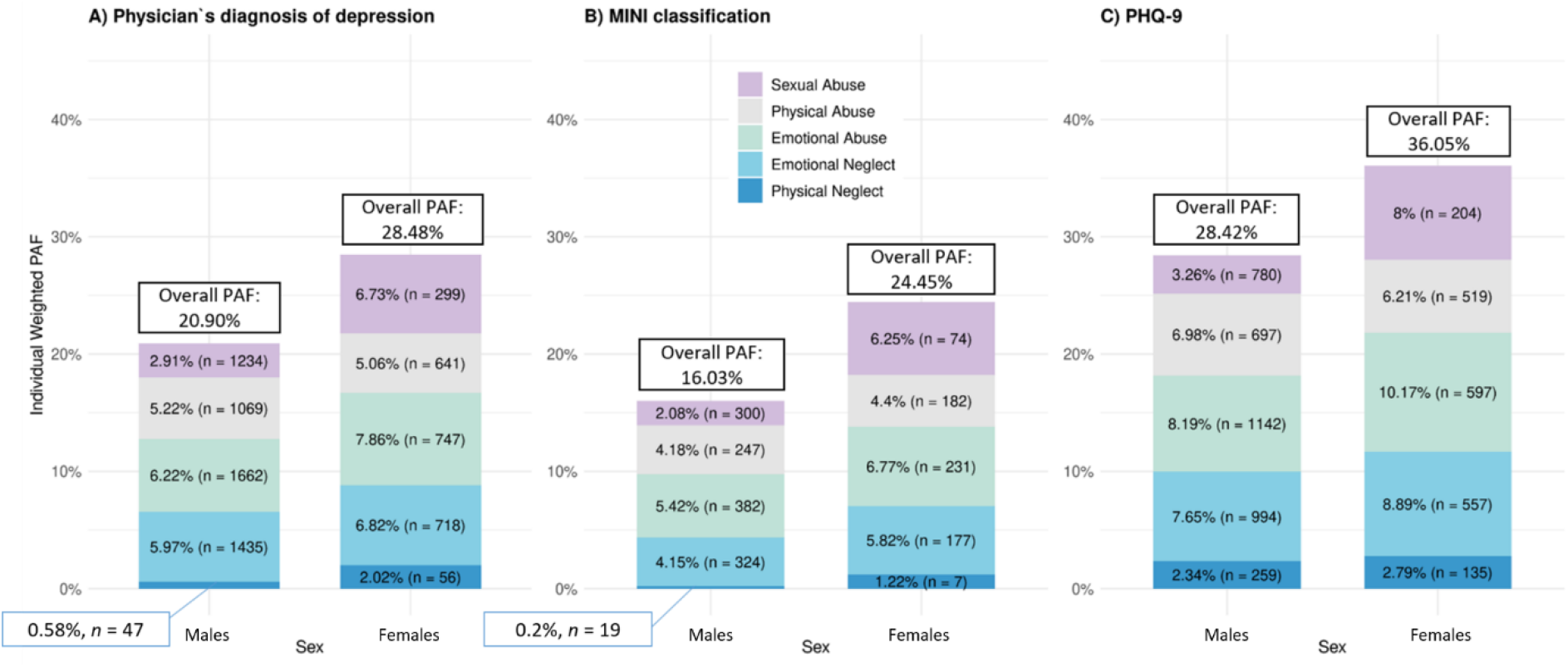
Population Attributable Fractions of Childhood Maltreatment and its Subtypes for Lifetime Depression and Current Depressive Symptoms Among Males and Females *Note:* PAF = Population Attributable Fraction. N_*males*_ = 80,519. N_*females*_ = 78,526. Sums of individual PAF estimates may differ slightly from overall PAF estimates due to rounding.

For MINI classification, sexual abuse (PAF_females_=6.3%; PAF_males_=2.1%) and emotional neglect (PAF_females_=5.8%; PAF_males_=4.2%) were associated with significantly higher PAF estimates in females than males (all p_adj_ < 0.007).

For current depressive symptoms, the overall PAF across the five maltreatment subtypes was 33.4% (95% CI [32.2%, 34.6%]), with individual subtype estimates ranging from 2.6% to 9.6%. The highest PAFs were observed for emotional abuse (PAF = 9.6%) and neglect (PAF = 8.4%, Figure 3).

The overall PAF for current depressive symptoms was significantly greater for females than males (p_adj_ < 0.005): In females, 36.1% (95% CI [34.5%, 37.7%]) of current depression were estimated to be associated with the five subtypes of maltreatment, in males only 28.4% (95% CI [26.7%, 30.2%]).

For current depression, higher PAFs in females than in males were observed for emotional (PAF_females_=10.2%; PAF_males_=8.2%, p_adj_ < 0.005) and sexual (PAF_females_= 8.0%; PAF_males_=3.3%, p_adj_ < 0.005) abuse, whereas for emotional neglect and physical subtypes, no significant sex difference was found (all p_adj_ > 0.05). Comparisons between the sex-stratified PAFs are presented in Supplementary Table S21. The PAFs calculated with Miettinen’s formula were identical to those calculated with Levin’s formula (see also [68]).

Sensitivity analyses using the joint-exposure approach yielded lower overall PAF estimates, although the overall pattern of findings remained unchanged. The PAF estimates across the complete sample were 22.8% (95% CI [22.0%, 23.7%]) for physician’s diagnosis, 19.2% (95% CI [18.2%, 21.0%]) for MINI classification, and 29.0% (95% CI [27.8%, 30.2%]) for current depressive symptoms, corresponding to a 9%-13% lower PAF observed with the joint-exposure approach. Additionally, females showed higher overall PAFs than males across all outcomes (physician’s diagnosis: 24.0% vs. 19.2%; MINI classification: 21.2% vs. 16.5%; current depressive symptoms: 30.4% vs. 26.1%; all p_adj_ < 0.001, Table S22), with PAFs being consistently lower with the joint-exposure approach except for the MINI classification in males. These findings indicate that although the estimated magnitude of the PAFs varies depending on the approach used to account for correlated childhood maltreatment exposures, the conclusions regarding the relative burden of childhood maltreatment and the observed sex differences were robust. The other sensitivity analyses adjusting for SES and in the L2 subset yielded comparable results to the main analyses (Tables S23–S29).

### Disparity Decomposition of Sex Differences in Depression by Childhood Maltreatment

Model-based disparity decomposition indicated that childhood maltreatment accounted for approximately one-fifth of the observed female excess in depression (physician’s diagnosis: 19.2%, MINI classification: 21.8%, PHQ-9: 22.4%). Most of this contribution was associated with females’ higher prevalence and joint distribution of childhood maltreatment (23.3–37.0% of the observed disparity), whereas sex-specific differences in the associations between maltreatment and depression contributed little and slightly reduced the overall disparity (−4.2% to −14.6%). These findings suggest that higher exposure to childhood maltreatment, rather than greater susceptibility to its depressive effects, accounted for most of the maltreatment-related component of the sex gap in depression (Table S30, Figure S4).

## Discussion

This study investigated sex-specific associations between childhood maltreatment and both lifetime and current depression in a large population-based cohort (NAKO). While depression was substantially more prevalent in females, sex differences in childhood maltreatment varied by subtype. Childhood maltreatment was associated with increased depression risk in both sexes, with subtype-specific associations varying by sex and a greater overall contribution to depression rates observed in females.

Across both sexes, *any maltreatment* and the single maltreatment subtypes showed ORs of >2 for both depression measures, with emotional subtypes - and for males, also sexual abuse - showing the strongest associations (OR > 3). Physical neglect showed substantially lower associations with depression (all OR < 1.5; and non-significant for physician’s diagnosis in males, and for MINI overall and in males), consistent with previous analyses of the NAKO dataset [18,19].

*Any maltreatment* showed a significant interaction with sex for physician’s diagnosis only, with a stronger association in females, which is in line with previous studies, showing that females are more prone to develop lifetime depression than males after childhood maltreatment [28,29]. Similarly, sex interactions of maltreatment subtypes were only observed for physician’s diagnosis, with stronger associations for physical subtypes in females and for sexual abuse in males. These findings have been reported in a recent article based on the NAKO dataset, examining lifetime depression [19]. The outcome-specific pattern for physician’s diagnosis may partly reflect differences in ascertainment rather than etiological mechanisms alone, as diagnoses depend on help-seeking behavior, access to care, and diagnostic practices, which are known to vary by sex, whereas self-reported symptoms may be less influenced by such factors [69,70]. While a meta-analysis from Gallo et al. reports consistent, but non-significant sex differences for physical abuse, stronger, but also non-significant associations between sexual abuse and depression in females than males were found [9]. This may reflect differences in outcome definitions or sample composition across studies. In the present study, a possible explanation for the stronger association with sexual abuse in males may be underreporting due to stigma, shame, and gender stereotypes, which might intensify its psychological impact [4]. In contrast, the weaker association between physical abuse and neglect and lifetime depression in males may reflect their lower tendency to perceive such events as traumatic [8]. Males are often socialized to be tough, resilient, and independent, and tend to perceive life events as more controllable [8,71]. Sex-specific social norms may promote more adaptive, problem-focused coping in males, but also externalizing behaviours (e.g., aggression or substance abuse). In contrast, females may be more prone to emotion-focused coping or internalizing responses [7,8,71].

As the results from this study and the existing evidence from other studies on the sex-specific impact of the subdimensions remain inconsistent, conclusions should be drawn with caution. While the main analyses were based on established CTS cut-offs commonly used in the literature, the sensitivity analyses indicated that the associations between childhood trauma and depression are not always adequately captured by a binary classification. Future research may therefore benefit from modeling the original CTS response categories to better characterize dose–response patterns and identify nuanced sex-specific associations across levels of trauma exposure. Given that the NAKO comprises the largest sample among these studies, the significant sex-specific associations observed for three of five subcategories on physician’s diagnosis indicate that sex plays a role in the associations.

The PAFs indicated that a notable share of depression occurrences in the NAKO sample is associated with childhood trauma, with the highest estimates for emotional subtypes. Sex-stratified PAFs revealed that, consistent with previous articles, a significantly higher proportion of depression cases in females were associated with childhood maltreatment than in males [40,43,44], with nearly all subtypes more strongly associated with depression in females, particularly the emotional subtypes and sexual abuse. The findings highlight that, in addition to the strength of the observed associations, the prevalence of the subtypes substantially contributes to the PAFs. For instance, although sexual abuse had a stronger association with physician’s diagnosis in males (OR_males_ ≈ 3.20, OR_females_ ≈ 2.65), it was about 3.5 times more prevalent among females, therefore explaining proportionally more of female depression cases. This offers a potential explanation of the stronger association of sexual abuse and depression in males, as they appear to have less male counterparts with comparable experiences and may experience stronger stigmatisation. Additionally, males were found to disclose sexual abuse less often, suggesting a high number of undetected cases. Given the 2:1 female-to-male ratio in overall depression prevalence, close to an estimated five out of six depression cases attributable to sexual abuse were observed in females. The disparity decomposition indicates that sex differences in childhood maltreatment prevalence and effects contribute to a meaningful but moderate proportion of the higher depression prevalence in females for the investigated depression outcomes. Nevertheless, the majority of the sex differences in depression remained independent of childhood maltreatment, highlighting that additional biological, social, or sex-specific mechanisms likely contribute to higher depression risk in females. Importantly, the disparity was largely driven by sex differences in the prevalence of childhood maltreatment subtypes rather than by sex-specific differences in their associations with depression.

Despite the observed sex differences in the present study, the results demonstrate that childhood trauma significantly influences and contributes to depression in both sexes. Most maltreatment subtypes show substantial associations with depression in both sexes (OR > 2); except for physical neglect. The PAF models indicate that childhood maltreatment accounted for 20.9% to 36.1% of NAKO depression cases, corresponding to a large share of NAKO depression occurrences in both sexes. The lower end of the PAF range is consistent with a recent study by Grummitt et al. estimating that approximately 21% of depression cases in Australia were attributable to childhood maltreatment [72]. Together, these findings suggest that childhood maltreatment represents an important shared pathway to depression in both sexes, contributing proportionally—and even more so in absolute numbers—to the population burden of depression among females than among males.

The present study has substantial strengths. It was carried out using data from the largest available dataset with CTS data from Germany to our knowledge. This allowed analyses with high statistical power [73], and the comprehensive investigations of sex-specific associations, and the investigation of childhood maltreatment subtypes and different depression definitions. Calculating PAFs not only takes into account both the association between childhood maltreatment and depression and the prevalence of the risk factors, but also the interrelatedness of the maltreatment subtypes [33], offering a broader understanding of the potential reduction in depression cases if exposure to childhood maltreatment were eliminated.

This study has several limitations. First, the retrospective self-report measures (i.e., CTS, physician’s diagnosis, MINI) [74] are susceptible to biases, particularly for sensitive information such as childhood maltreatment which some individuals might be unwilling to disclose [50]. Moreover, individuals with (current) depression may recall childhood experiences more negatively [74]. Additionally, the CTS measures each subtype with a single item, which may limit the depth and specificity compared to the full CTQ or impact effect estimates [75]. Furthermore, information on the timing and severity of childhood maltreatment was not recorded [16,18]. Notably, physical neglect was the only subtype with a non-significant association with the MINI across sexes, and lifetime depression in males. This may result from low item sensitivity, meaning that the item insufficiently captures its breadth and severity and leading to underreporting and attenuation of the association [51]. Additionally, prior findings indicate that the physical neglect item shows inconsistent reliability, comparatively low correlations with related measures, and unsatisfactory internal consistency, suggesting it may not be well differentiated from other subscales [49,76]. Nonetheless, the five-item CTS is an economical approach, suitable for large-scale cohorts like the NAKO. Furthermore, participants over 40 years of age were oversampled, which limited the sample’s representativeness. Although the response (∼ 15.6%) is relatively high for a large-scale cohort study [45,77], selection bias remains relevant, as participants with low SES, who are more often affected by childhood maltreatment [50] and depression [23], and individuals with severe depression are likely underrepresented in the sample. Further, primary models were only adjusted for age, age² and education, leaving residual confounding likely. A recent meta-analysis of quasi-experimental studies found that associations between childhood maltreatment and mental health outcomes were substantially attenuated after rigorous adjustment for shared genetic and environmental confounding, although a significant association remained, suggesting that both causal effects and residual confounding contribute to observed associations[78]. Therefore, the present findings should not be interpreted as establishing causality. Accordingly, PAFs represent model-based estimates under specific assumptions rather than true causal proportions of depression cases. Moreover, non-binary sex has not been assessed in the NAKO, potentially masking effects specific to intersex individuals. Furthermore, sex information derived from the residency register reflects administrative sex rather than biological sex characteristics or gender identity. Until 2018, administrative classification allowed only binary biological sex (female/male). A third category (diverse) was introduced in 2018 [79]. In 2024, Germany passed a law that simplifies applications to change registered gender entries [80]. Changes to registered sex prior to this law were very rare, therefore, it can be assumed that the administrative sex recorded in the NAKO between 2015 and 2019 largely corresponds to biological sex at birth. Using gender identity instead of biological sex might have yielded different effects specific to a female, male and diverse (i.e., transgender and non-binary) gender identity. A recent review by Arena et al. has found that transgender individuals more frequently reported adverse childhood experiences than cisgender individuals, highlighting the need to investigate transgender-specific effects of childhood maltreatment on mental health [81].

Given the cross-sectional design of the present analysis, future longitudinal NAKO data [17] will provide more valuable insights into the childhood maltreatment-depression relationship and its public health implications. Further research should also integrate biological factors, social networks and socio-behavioral measures that may influence subtype-specific and sex-specific associations [9,11,71,82].

## Conclusion

The findings highlight the significant impact of childhood maltreatment and its subtypes on both lifetime and current depression, with sex-specific associations observed only for physician’s diagnosis. To our knowledge, this is among the first large population-based studies differentiating subtype-specific childhood maltreatment associations with both lifetime and current depression. A substantial share of depression cases, especially among females, was associated with childhood maltreatment. Sex-specific prevention and coping strategies could help reduce overall depression prevalence [83].

## Statements

## Data Availability

All data produced in the present study are available upon reasonable request to the authors at the NAKO transfer hub (https://transfer.nako.de/)

## Acknowledgement

We thank all participants, the complete staff at the NAKO study centers, the data management and integration center, and the NAKO head office who enabled the conduction of the study and made the collection of all data possible.

## Statement of Ethics

The present study was initially approved by the ethical committee of the Bavarian State Medical Association (Nr. 13023). Subsequently, all local ethical committees approved the study.

Written consent was obtained from all participants before participation.

## Author Contributions

MPV, ALK, and FS contributed to conceptualization, with input from JF, JKK, LZ, ES, KB, JM, and SHW. Methodology was developed by MPV, ALK, and FS, with contributions from JF, JKK, LZ, ES, KB, JM, and SHW. MPV and ALK performed the formal analyses, with methodological advice from JF and IR. Data investigation was carried out by RM, ML, PB, LK, TK, CMF, SRH, HG, BB, HB, NO, VH, TP, and KB, who also contributed to data curation. MPV and ALK drafted the original manuscript. JCF, DFS, and FS contributed to manuscript review and editing, along with all other authors. FS provided supervision.

All authors contributed to interpretation of the results, critically revised the manuscript for important intellectual content, and approved the final version for submission.

## Data Availability Statement

The datasets analyzed in the current study are not publicly available due to privacy restrictions but can be requested through the NAKO transfer hub (https://transfer.nako.de/). Data access for this study was granted under application number NAKO-679.

## Code Availability Statement

All code used for data processing, analysis, and figure generation in this study is publicly available at https://doi.org/10.17605/OSF.IO/K48DZ.

