## Supplemental Figure 4 for "Examining the Contribution of Childhood Maltreatment to the Sex Gap in Depression: Insights from the German National Cohort (NAKO)"

**Figure S4.** Disparity decomposition of sex differences in depression by childhood maltreatment.

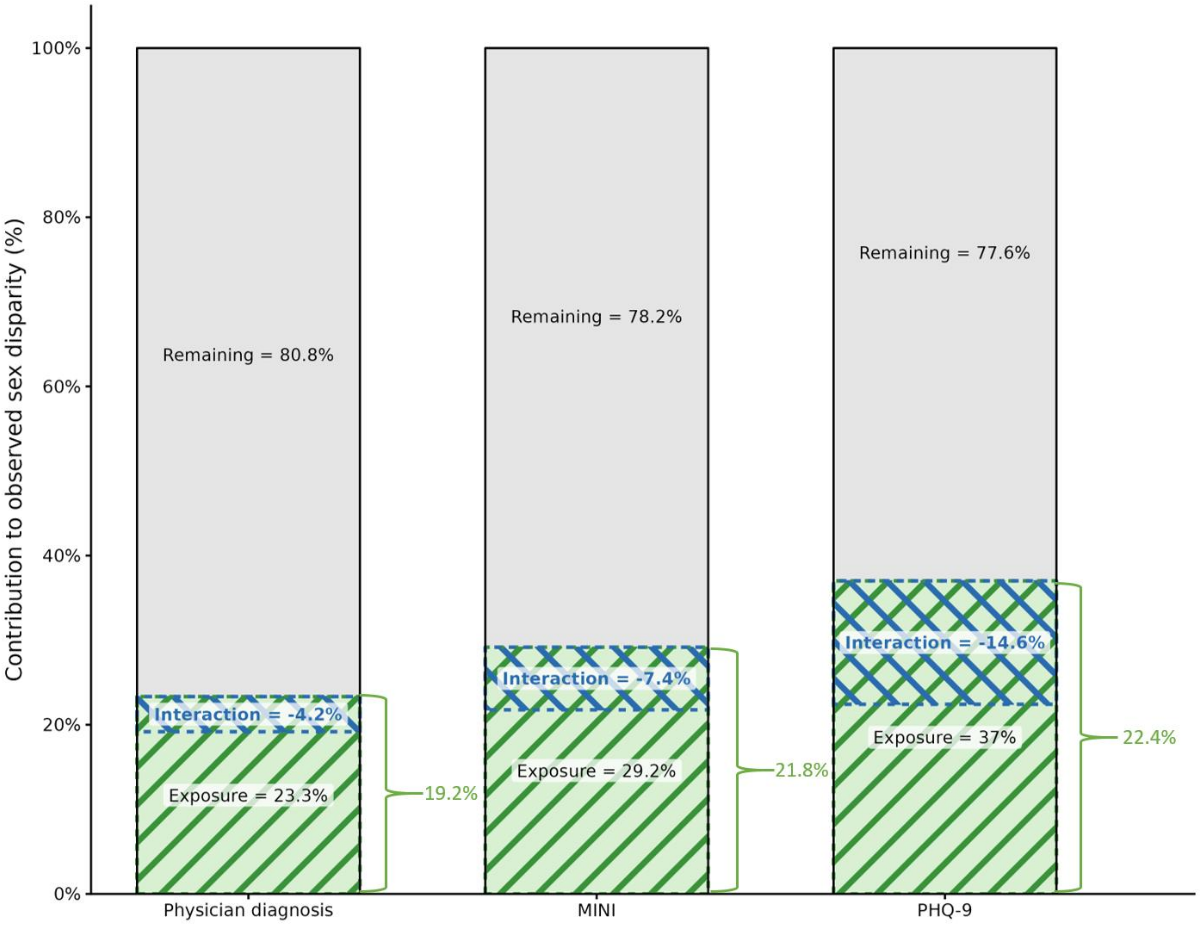


*Note.* MINI = Mini International Neuropsychiatric Interview. PHQ-9 = Patient Health Questionnaire. Exposure = the proportion of the observed disparity attributable to sex differences in the prevalence of childhood maltreatment. Interaction = the reduction in the exposure component due to sex differences in the effect of childhood maltreatment on depression. Because the interaction component is negative, it is displayed as an overlap within the exposure component. Brackets indicate the total proportion of the observed disparity accounted for by childhood maltreatment (exposure + interaction). Remaining = remaining unexplained disparity. Minor discrepancies between the displayed component values and the total explained proportion are due to rounding.
