## Supplementary Material for "Examining the Contribution of Childhood Maltreatment to the Sex Gap in Depression: Insights from the German National Cohort (NAKO)"

### Table S1. *Frequency Distribution of Response Categories to Childhood Trauma Screener Items* (N = 159,045)

|  | Mean | SD | Never | Rarely | Sometimes | Often | Very often | Maltreated |
| --- | --- | --- | --- | --- | --- | --- | --- | --- |
| Sexual Abuse: Someone molested me (sexually). | 1.10 | 0.45 | 93.8% | 3.3% | 2.1% | 0.5% | 0.4% | 6.0% |
| Physical Abuse: People in my family hit me so hard that it left me with bruises or marks. | 1.31 | 0.74 | 80.7% | 11.1% | 5.4% | 1.8% | 1.0% | 8.0% |
| Emotional Abuse: I felt that someone in my family hated me | 1.30 | 0.78 | 83.8% | 8.0% | 4.6% | 2.3% | 1.4% | 8.0% |
| Emotional Neglect:  I felt loved. (R) | 1.77 | 0.96 | 49.1% | 34.2% | 9.1% | 5.9% | 1.8% | 7.4% |
| Physical Neglect: There was someone to take me to the doctor if I needed it. (R) | 1.79 | 1.15 | 58.9% | 18.9% | 12.3% | 4.7% | 5.2% | 9.7% |

*Note*. Responses indicating ‘moderate/severe trauma’ are shaded in grey, according to Glaesmer et al. (2013). (R) = Reversed coding.

### Table S2. *Data Availability and Missingness for Childhood Maltreatment and Depression Measures.*

|  | Valid n | % Valid | Missings | % Missings |
| --- | --- | --- | --- | --- |
| Total sample | 204.725 |  |  |  |
| CTS *any maltreatment* | 172.038 | 84.03% | 32.687 | 15.97% |
| CTS Sexual Abuse | 172.081 | 84.05% | 32.644 | 15.95% |
| CTS Physical Abuse | 172.077 | 84.05% | 32.648 | 15.95% |
| CTS Emotional Abuse | 172.075 | 84.05% | 32.650 | 15.95% |
| CTS Emotional Neglect | 172.084 | 84.06% | 32.641 | 15.94% |
| CTS Physical Neglect | 172.061 | 84.04% | 32.664 | 15.96% |
| Physician’s Diagnosis | 203.152 | 99.23% | 1.573 | 0.77% |
| PHQ-9 | 189.340 | 92.49% | 15.385 | 7.51% |
| L2 participants | 60.418 |  |  |  |
| MINI Classification | 57.803 | 95.67% | 2.615 | 4.33% |

*Note.* CTS = Childhood Trauma Screener. MINI = Mini International Neuropsychiatric Interview. PHQ-9 = Patient Health Questionnaire. L2 = Level-2. Valid n and % Valid refer to the number and percentage of participants with available data for the respective variable. Missing values include all participants with unavailable, incomplete, or non-interpretable data. Percentages were calculated relative to the total sample size (N = 204,725) or, for MINI classifications, relative to the Level-2 diagnostic subsample (n = 60,418).

### Table S3. *Summarized Results of the Multiple Binary Logistic Regression Analysis with Depression Measures as Outcome Variables in the Complete Sample*

| Variables | *OR* | *OR 95%*  *CI lower* | *OR 95%*  *CI upper* | *Estimate* | *Std. Error* | *z-Value* | *P_adj_* |
| --- | --- | --- | --- | --- | --- | --- | --- |
| **Physician’s diagnosis** |  |  |  |  |  |  |  |
| CTS | 2.45 | 2.38 | 2.53 | 0.90 | 0.02 | 58.76 | < .001** |
| CTS SA | 3.16 | 3.02 | 3.31 | 1.15 | 0.02 | 49.18 | < .001** |
| CTS PA | 2.48 | 2.38 | 2.59 | 0.91 | 0.02 | 42.31 | < .001** |
| CTS EA | 3.50 | 3.36 | 3.65 | 1.25 | 0.02 | 60.81 | < .001** |
| CTS EN | 3.33 | 3.19 | 3.47 | 1.20 | 0.02 | 56.22 | < .001** |
| CTS PN | 1.23 | 1.18 | 1.29 | 0.21 | 0.02 | 9.10 | < .001** |
| **MINI** |  |  |  |  |  |  |  |
| CTS | 2.30 | 2.18 | 2.43 | 0.83 | 0.03 | 29.38 | < .001** |
| CTS SA | 2.89 | 2.64 | 3.16 | 1.06 | 0.05 | 23.09 | < .001** |
| CTS PA | 2.33 | 2.15 | 2.53 | 0.85 | 0.04 | 20.55 | < .001** |
| CTS EA | 3.32 | 3.07 | 3.58 | 1.20 | 0.04 | 30.28 | < .001** |
| CTS EN | 2.93 | 2.70 | 3.18 | 1.08 | 0.04 | 25.70 | < .001** |
| CTS PN | 1.12 | 1.03 | 1.23 | 0.12 | 0.04 | 2.66 | .141 |
| **PHQ-9** |  |  |  |  |  |  |  |
| CTS | 2.90 | 2.79 | 3.02 | 1.06 | 0.02 | 53.32 | < .001** |
| CTS SA | 3.08 | 2.91 | 3.26 | 1.13 | 0.03 | 39.27 | < .001** |
| CTS PA | 2.82 | 2.68 | 2.97 | 1.04 | 0.03 | 39.10 | < .001** |
| CTS EA | 3.97 | 3.79 | 4.17 | 1.38 | 0.02 | 56.71 | < .001** |
| CTS EN | 3.79 | 3.60 | 3.99 | 1.33 | 0.03 | 51.08 | < .001** |
| CTS PN | 1.41 | 1.33 | 1.49 | 0.34 | 0.03 | 11.44 | < .001** |

*Note.* OR = Odds Ratio. CTS = Childhood Trauma Screener. SA = Sexual Abuse. PA = Physical Abuse. EA = Emotional Abuse. EN = Emotional Neglect. PN = Physical Neglect. MINI = Mini International Neuropsychiatric Interview. PHQ-9 = Patient Health Questionnaire. Odds Ratios with 95% confidence intervals are presented for categorical measures. As reference group, participants who reported no/low trauma were selected. P_adj_ = adjusted *p*-values. ** *p_adj_* < .001 * *p_adj_* < .05. Adjusted *p*-values exceeding 1 are reported as *p* = 1.000.

### Table S4. *Summarized Results of the Sex-Stratified Binary Logistic Regression Analysis with Depression Measures as Outcome Variables for Females*

| Variables | *OR* | *OR 95%*  *CI lower* | *OR 95%*  *CI upper* | *Estimate* | *Std. Error* | *z-Value* | *P_adj_* |
| --- | --- | --- | --- | --- | --- | --- | --- |
| **Physician’s diagnosis** |  |  |  |  |  |  |  |
| CTS | 2.53 | 2.43 | 2.62 | 0.93 | 0.02 | 47.33 | < .001** |
| CTS SA | 2.61 | 2.48 | 2.75 | 0.96 | 0.03 | 36.14 | < .001** |
| CTS PA | 2.74 | 2.59 | 2.90 | 1.01 | 0.03 | 34.99 | < .001** |
| CTS EA | 3.31 | 3.15 | 3.49 | 1.20 | 0.03 | 46.49 | < .001** |
| CTS EN | 3.37 | 3.19 | 3.56 | 1.21 | 0.03 | 44.04 | < .001** |
| CTS PN | 1.36 | 1.28 | 1.44 | 0.31 | 0.03 | 10.44 | < .001** |
| **MINI** |  |  |  |  |  |  |  |
| CTS | 2.36 | 2.19 | 2.54 | 0.86 | 0.04 | 22.55 | < .001** |
| CTS SA | 2.54 | 2.29 | 2.81 | 0.93 | 0.05 | 17.65 | < .001** |
| CTS PA | 2.51 | 2.25 | 2.81 | 0.92 | 0.06 | 16.12 | < .001** |
| CTS EA | 3.06 | 2.76 | 3.38 | 1.12 | 0.05 | 21.78 | < .001** |
| CTS EN | 3.01 | 2.70 | 3.35 | 1.10 | 0.06 | 20.00 | < .001** |
| CTS PN | 1.21 | 1.08 | 1.36 | 0.19 | 0.06 | 3.23 | .045* |
| **PHQ-9** |  |  |  |  |  |  |  |
| CTS | 2.88 | 2.73 | 3.03 | 1.06 | 0.03 | 40.85 | < .001** |
| CTS SA | 2.72 | 2.55 | 2.90 | 1.00 | 0.03 | 30.46 | < .001** |
| CTS PA | 2.92 | 2.72 | 3.12 | 1.07 | 0.04 | 30.40 | < .001** |
| CTS EA | 3.73 | 3.51 | 3.96 | 1.32 | 0.03 | 43.12 | < .001** |
| CTS EN | 3.79 | 3.55 | 4.05 | 1.33 | 0.03 | 39.97 | < .001** |
| CTS PN | 1.47 | 1.36 | 1.58 | 0.38 | 0.04 | 9.97 | < .001** |

*Note.* OR = Odds Ratio. CTS = Childhood Trauma Screener. SA = Sexual Abuse. PA = Physical Abuse. EA = Emotional Abuse. EN = Emotional Neglect. PN = Physical Neglect. MINI = Mini International Neuropsychiatric Interview. PHQ-9 = Patient Health Questionnaire. Odds Ratios with 95% confidence intervals are presented for categorical measures. As reference group, participants who reported no/low trauma were selected. P_adj_ = adjusted *p*-values. ** *p_adj_* < .001 * *p_adj_* < .05. Adjusted *p*-values exceeding 1 are reported as *p* = 1.000.

### *Table S5. Summarized Results of the Sex-Stratified Binary Logistic Regression Analysis with Depression Measures as Outcome Variables for Males*

| Variables | *OR* | *OR 95%*  *CI lower* | *OR 95%*  *CI upper* | *Estimate* | *Std. Error* | *z-Value* | *P_adj_* |
| --- | --- | --- | --- | --- | --- | --- | --- |
| **Physician’s diagnosis** |  |  |  |  |  |  |  |
| CTS | 2.19 | 2.09 | 2.30 | 0.79 | 0.02 | 31.50 | < .001** |
| CTS SA | 3.23 | 2.91 | 3.57 | 1.17 | 0.05 | 22.53 | < .001** |
| CTS PA | 2.36 | 2.21 | 2.52 | 0.86 | 0.03 | 25.84 | < .001** |
| CTS EA | 3.37 | 3.14 | 3.60 | 1.21 | 0.04 | 34.55 | < .001** |
| CTS EN | 3.11 | 2.91 | 3.33 | 1.14 | 0.03 | 32.60 | < .001** |
| CTS PN | 1.08 | 1.00 | 1.16 | 0.08 | 0.04 | 2.09 | 1.000 |
| **MINI** |  |  |  |  |  |  |  |
| CTS | 2.13 | 1.95 | 2.31 | 0.75 | 0.04 | 17.48 | < .001** |
| CTS SA | 2.80 | 2.30 | 3.39 | 1.03 | 0.10 | 10.35 | < .001** |
| CTS PA | 2.23 | 1.97 | 2.50 | 0.80 | 0.06 | 13.19 | < .001** |
| CTS EA | 3.40 | 3.00 | 3.84 | 1.22 | 0.06 | 19.44 | < .001** |
| CTS EN | 2.66 | 2.34 | 3.03 | 0.98 | 0.07 | 14.91 | < .001** |
| CTS PN | 1.03 | 0.90 | 1.18 | 0.03 | 0.07 | 0.44 | 1.000 |
| **PHQ-9** |  |  |  |  |  |  |  |
| CTS | 2.80 | 2.63 | 2.97 | 1.03 | 0.03 | 32.49 | < .001** |
| CTS SA | 3.28 | 2.90 | 3.71 | 1.19 | 0.06 | 19.05 | < .001** |
| CTS PA | 2.81 | 2.59 | 3.04 | 1.03 | 0.04 | 25.29 | < .001** |
| CTS EA | 4.04 | 3.73 | 4.37 | 1.40 | 0.04 | 34.42 | < .001** |
| CTS EN | 3.62 | 3.33 | 3.94 | 1.29 | 0.04 | 30.48 | < .001** |
| CTS PN | 1.33 | 1.22 | 1.46 | 0.29 | 0.05 | 6.11 | < .001** |

*Note.* OR = Odds Ratio. CTS = Childhood Trauma Screener. SA = Sexual Abuse. PA = Physical Abuse. EA = Emotional Abuse. EN = Emotional Neglect. PN = Physical Neglect. MINI = Mini International Neuropsychiatric Interview. PHQ-9 = Patient Health Questionnaire. Odds Ratios with 95% confidence intervals are presented for categorical measures. As reference group, participants who reported no/low trauma were selected. P_adj_ = adjusted *p*-values. ** *p_adj_* < .001 * *p_adj_* < .05. Adjusted *p*-values exceeding 1 are reported as *p* = 1.000.

### Table S6. *Summarized Results of the Multiple Binary Logistic Regression Analysis with Sex Interactions with Depression Measures as Outcome Variables*

| Variables | *OR* | *OR 95%*  *CI lower* | *OR 95%*  *CI upper* | *Estimate* | *Std. Error* | *z-Value* | *P_adj_* |
| --- | --- | --- | --- | --- | --- | --- | --- |
| **Physician’s diagnosis** |  |  |  |  |  |  |  |
| CTS*sex | 1.14 | 1.07 | 1.21 | 0.13 | 0.03 | 4.23 | < .001** |
| SA*sex | 0.80 | 0.72 | 0.90 | -0.22 | 0.06 | -3.78 | .003* |
| PA*sex | 1.15 | 1.06 | 1.25 | 0.14 | 0.04 | 3.21 | .024* |
| EA*sex | 0.97 | 0.89 | 1.06 | -0.03 | 0.04 | -0.70 | 1.000 |
| EN*sex | 1.06 | 0.97 | 1.16 | 0.06 | 0.04 | 1.38 | 1.000 |
| PN*sex | 1.23 | 1.12 | 1.35 | 0.21 | 0.05 | 4.41 | < .001** |
| **MINI** |  |  |  |  |  |  |  |
| CTS*sex | 1.11 | 0.99 | 1.24 | 0.10 | 0.06 | 1.79 | 1.000 |
| SA*sex | 0.90 | 0.72 | 1.12 | -0.10 | 0.11 | -0.93 | 1.000 |
| PA*sex | 1.13 | 0.96 | 1.32 | 0.12 | 0.08 | 1.43 | 1.000 |
| EA*sex | 0.87 | 0.74 | 1.02 | -0.14 | 0.08 | -1.74 | 1.000 |
| EN*sex | 1.11 | 0.94 | 1.31 | 0.11 | 0.08 | 1.25 | 1.000 |
| PN*sex | 1.17 | 0.98 | 1.39 | 0.16 | 0.09 | 1.78 | 1.000 |
| **PHQ-9** |  |  |  |  |  |  |  |
| CTS*sex | 1.04 | 0.96 | 1.12 | 0.04 | 0.04 | 0.90 | 1.000 |
| SA*sex | 0.83 | 0.72 | 0.95 | -0.19 | 0.07 | -2.68 | .132 |
| PA*sex | 1.04 | 0.94 | 1.16 | 0.04 | 0.05 | 0.82 | 1.000 |
| EA*sex | 0.92 | 0.83 | 1.01 | -0.09 | 0.05 | -1.74 | 1.000 |
| EN*sex | 1.04 | 0.94 | 1.16 | 0.04 | 0.05 | 0.83 | 1.000 |
| PN*sex | 1.09 | 0.97 | 1.23 | 0.09 | 0.06 | 1.51 | 1.000 |

*Note.* OR = Odds Ratio. CTS = Childhood Trauma Screener. SA = Sexual Abuse. PA = Physical Abuse. EA = Emotional Abuse. EN = Emotional Neglect. PN = Physical Neglect. MINI = Mini International Neuropsychiatric Interview. PHQ-9 = Patient Health Questionnaire. Odds Ratios with 95% confidence intervals are presented for categorical measures. As reference group, participants who reported no/low trauma were selected. P_adj_ = adjusted *p*-values. ** *p_adj_* < .001 * *p_adj_* < .05. Adjusted *p*-values exceeding 1 are reported as *p* = 1.000.

### Table S7. *Summarized Results of the Sensitivity Analysis with Depression Measures as Outcome Variables in the Complete Sample*

| Variables | *OR* | *OR 95%*  *CI lower* | *OR 95%*  *CI upper* | *Estimate* | *Std. Error* | *z-Value* | *P_adj_* |
| --- | --- | --- | --- | --- | --- | --- | --- |
| **Physician’s diagnosis** |  |  |  |  |  |  |  |
| CTS | 2.37 | 2.29 | 2.44 | 0.86 | 0.02 | 54.60 | < .001** |
| CTS SA | 3.07 | 2.92 | 3.21 | 1.12 | 0.02 | 46.38 | < .001** |
| CTS PA | 2.39 | 2.29 | 2.50 | 0.87 | 0.02 | 39.23 | < .001** |
| CTS EA | 3.34 | 3.20 | 3.48 | 1.21 | 0.02 | 56.57 | < .001** |
| CTS EN | 3.14 | 3.01 | 3.28 | 1.15 | 0.02 | 51.64 | < .001** |
| CTS PN | 1.18 | 1.12 | 1.23 | 0.16 | 0.02 | 6.82 | < .001** |
| **MINI** |  |  |  |  |  |  |  |
| CTS | 2.24 | 2.11 | 2.37 | 0.80 | 0.03 | 27.54 | < .001** |
| CTS SA | 2.89 | 2.64 | 3.16 | 1.06 | 0.05 | 23.09 | < .001** |
| CTS PA | 2.33 | 2.15 | 2.53 | 0.85 | 0.04 | 20.55 | < .001** |
| CTS EA | 3.22 | 2.97 | 3.48 | 1.17 | 0.04 | 28.63 | < .001** |
| CTS EN | 2.81 | 2.58 | 3.06 | 1.03 | 0.04 | 23.83 | < .001** |
| CTS PN | 1.09 | 1.00 | 1.19 | 0.09 | 0.05 | 1.91 | 1.000 |
| **PHQ-9** |  |  |  |  |  |  |  |
| CTS | 2.42 | 2.12 | 2.78 | 0.89 | 0.07 | 12.80 | < .001** |
| CTS SA | 2.93 | 2.76 | 3.10 | 1.07 | 0.03 | 36.08 | < .001** |
| CTS PA | 2.68 | 2.54 | 2.83 | 0.99 | 0.03 | 35.82 | < .001** |
| CTS EA | 3.71 | 3.53 | 3.90 | 1.31 | 0.03 | 51.77 | < .001** |
| CTS EN | 3.47 | 3.29 | 3.66 | 1.25 | 0.03 | 45.78 | < .001** |
| CTS PN | 1.31 | 1.23 | 1.39 | 0.27 | 0.03 | 8.76 | < .001** |

*Note.* Sensitivity analysis with relative income position as additional covariate. OR = Odds Ratio. CTS = Childhood Trauma Screener. SA = Sexual Abuse. PA = Physical Abuse. EA = Emotional Abuse. EN = Emotional Neglect. PN = Physical Neglect. MINI = Mini International Neuropsychiatric Interview. PHQ-9 = Patient Health Questionnaire. Odds Ratios with 95% confidence intervals are presented for categorical measures. As reference group, participants who reported no/low trauma were selected. P_adj_ = adjusted *p*-values. ** *p_adj_* < .001 * *p_adj_* < .05. Adjusted *p*-values exceeding 1 are reported as *p* = 1.000.

### Table S8. *Summarized Results of the Sensitivity Analysis with Depression Measures as Outcome Variables for Females*

| Variables | *OR* | *OR 95%*  *CI lower* | *OR 95%*  *CI upper* | *Estimate* | *Std. Error* | *z-Value* | *P_adj_* |
| --- | --- | --- | --- | --- | --- | --- | --- |
| **Physician’s diagnosis** |  |  |  |  |  |  |  |
| CTS | 2.46 | 2.36 | 2.56 | 0.90 | 0.02 | 44.41 | < .001** |
| CTS SA | 2.55 | 2.42 | 2.70 | 0.94 | 0.03 | 34.11 | < .001** |
| CTS PA | 2.64 | 2.49 | 2.80 | 0.97 | 0.03 | 32.55 | < .001** |
| CTS EA | 3.18 | 3.02 | 3.35 | 1.16 | 0.03 | 43.37 | < .001** |
| CTS EN | 3.20 | 3.02 | 3.38 | 1.16 | 0.03 | 40.61 | < .001** |
| CTS PN | 1.31 | 1.24 | 1.39 | 0.27 | 0.03 | 8.93 | < .001** |
| **MINI** |  |  |  |  |  |  |  |
| CTS | 2.30 | 2.13 | 2.49 | 0.83 | 0.04 | 21.26 | < .001** |
| CTS SA | 2.48 | 2.22 | 2.75 | 0.91 | 0.05 | 16.70 | < .001** |
| CTS PA | 2.46 | 2.19 | 2.76 | 0.90 | 0.06 | 15.29 | < .001** |
| CTS EA | 2.98 | 2.69 | 3.31 | 1.09 | 0.05 | 20.68 | < .001** |
| CTS EN | 2.89 | 2.58 | 3.23 | 1.06 | 0.06 | 18.59 | < .001** |
| CTS PN | 1.18 | 1.05 | 1.33 | 0.17 | 0.06 | 2.73 | .224 |
| **PHQ-9** |  |  |  |  |  |  |  |
| CTS | 2.75 | 2.61 | 2.90 | 1.01 | 0.03 | 37.68 | < .001** |
| CTS SA | 2.63 | 2.46 | 2.81 | 0.97 | 0.03 | 28.34 | < .001** |
| CTS PA | 2.79 | 2.60 | 3.00 | 1.03 | 0.04 | 28.11 | < .001** |
| CTS EA | 3.52 | 3.31 | 3.75 | 1.26 | 0.03 | 39.58 | < .001** |
| CTS EN | 3.50 | 3.27 | 3.75 | 1.25 | 0.03 | 36.07 | < .001** |
| CTS PN | 1.38 | 1.28 | 1.49 | 0.32 | 0.04 | 8.08 | < .001** |

*Note.* Sensitivity analysis with relative income position as additional covariate. OR = Odds Ratio. CTS = Childhood Trauma Screener. SA = Sexual Abuse. PA = Physical Abuse. EA = Emotional Abuse. EN = Emotional Neglect. PN = Physical Neglect. MINI = Mini International Neuropsychiatric Interview. PHQ-9 = Patient Health Questionnaire. Odds Ratios with 95% confidence intervals are presented for categorical measures. As reference group, participants who reported no/low trauma were selected. P_adj_ = adjusted *p*-values. ** *p_adj_* < .001 * *p_adj_* < .05. Adjusted *p*-values exceeding 1 are reported as *p* = 1.000.

### *Table S9. Summarized Results of the Sensitivity Analysis with Depression Measures as Outcome Variables for Males*

| Variables | *OR* | *OR 95%*  *CI lower* | *OR 95%*  *CI upper* | *Estimate* | *Std. Error* | *z-Value* | *P_adj_* |
| --- | --- | --- | --- | --- | --- | --- | --- |
| **Physician’s diagnosis** |  |  |  |  |  |  |  |
| CTS | 2.10 | 1.99 | 2.20 | 0.74 | 0.03 | 28.82 | < .001** |
| CTS SA | 3.09 | 2.78 | 3.43 | 1.13 | 0.05 | 21.16 | < .001** |
| CTS PA | 2.27 | 2.12 | 2.43 | 0.82 | 0.03 | 23.90 | < .001** |
| CTS EA | 3.19 | 2.97 | 3.43 | 1.16 | 0.04 | 32.03 | < .001** |
| CTS EN | 2.94 | 2.73 | 3.15 | 1.08 | 0.04 | 29.88 | < .001** |
| CTS PN | 1.02 | 0.94 | 1.10 | 0.02 | 0.04 | 0.49 | 1.000 |
| **MINI** |  |  |  |  |  |  |  |
| CTS | 2.06 | 1.88 | 2.24 | 0.72 | 0.04 | 16.24 | < .001** |
| CTS SA | 2.74 | 2.24 | 3.35 | 1.01 | 0.10 | 9.85 | < .001** |
| CTS PA | 2.20 | 1.94 | 2.48 | 0.79 | 0.06 | 12.67 | < .001** |
| CTS EA | 3.27 | 2.88 | 3.71 | 1.19 | 0.06 | 18.28 | < .001** |
| CTS EN | 2.55 | 2.23 | 2.91 | 0.94 | 0.07 | 13.79 | < .001** |
| CTS PN | 0.99 | 0.87 | 1.14 | -0.01 | 0.07 | -0.08 | 1.000 |
| **PHQ-9** |  |  |  |  |  |  |  |
| CTS | 2.60 | 2.43 | 2.77 | 0.95 | 0.03 | 29.06 | < .001** |
| CTS SA | 3.01 | 2.65 | 3.41 | 1.10 | 0.06 | 17.04 | < .001** |
| CTS PA | 2.64 | 2.43 | 2.87 | 0.97 | 0.04 | 22.87 | < .001** |
| CTS EA | 3.74 | 3.45 | 4.07 | 1.32 | 0.04 | 31.27 | < .001** |
| CTS EN | 3.29 | 3.02 | 3.59 | 1.19 | 0.04 | 27.09 | < .001** |
| CTS PN | 1.23 | 1.12 | 1.35 | 0.21 | 0.05 | 4.24 | .001* |

*Note.* Sensitivity analysis with relative income position as additional covariate. OR = Odds Ratio. CTS = Childhood Trauma Screener. SA = Sexual Abuse. PA = Physical Abuse. EA = Emotional Abuse. EN = Emotional Neglect. PN = Physical Neglect. MINI = Mini International Neuropsychiatric Interview. PHQ-9 = Patient Health Questionnaire. Odds Ratios with 95% confidence intervals are presented for categorical measures. As reference group, participants who reported no/low trauma were selected. P_adj_ = adjusted *p*-values. ** *p_adj_* < .001 * *p_adj_* < .05. Adjusted *p*-values exceeding 1 are reported as *p* = 1.000.

### Table S10. *Summarized Results of the Sensitivity Analysis with Sex Interactions with Depression Measures as Outcome Variables*

| Variables | *OR* | *OR 95%*  *CI lower* | *OR 95%*  *CI upper* | *Estimate* | *Std. Error* | *z-Value* | *P_adj_* |
| --- | --- | --- | --- | --- | --- | --- | --- |
| **Physician’s diagnosis** |  |  |  |  |  |  |  |
| CTS*sex | 1.16 | 1.09 | 1.24 | 0.15 | 0.03 | 4.61 | < .001** |
| SA*sex | 0.82 | 0.73 | 0.92 | -0.20 | 0.06 | -3.40 | .013* |
| PA*sex | 1.15 | 1.05 | 1.26 | 0.14 | 0.05 | 3.13 | .032* |
| EA*sex | 0.98 | 0.90 | 1.07 | -0.02 | 0.04 | -0.46 | 1.000 |
| EN*sex | 1.07 | 0.98 | 1.17 | 0.07 | 0.05 | 1.45 | 1.000 |
| PN*sex | 1.26 | 1.15 | 1.39 | 0.23 | 0.05 | 4.74 | < .001** |
| **MINI** |  |  |  |  |  |  |  |
| CTS*sex | 1.12 | 1.00 | 1.25 | 0.11 | 0.06 | 1.93 | .981 |
| SA*sex | 0.90 | 0.72 | 1.13 | -0.11 | 0.12 | -0.95 | 1.000 |
| PA*sex | 1.12 | 0.95 | 1.32 | 0.11 | 0.08 | 1.32 | 1.000 |
| EA*sex | 0.88 | 0.75 | 1.04 | -0.13 | 0.08 | -1.54 | 1.000 |
| EN*sex | 1.12 | 0.94 | 1.33 | 0.11 | 0.09 | 1.26 | 1.000 |
| PN*sex | 1.19 | 0.99 | 1.42 | 0.17 | 0.09 | 1.88 | 1.000 |
| **PHQ-9** |  |  |  |  |  |  |  |
| CTS*sex | 1.07 | 0.98 | 1.16 | 0.07 | 0.04 | 1.58 | 1.000 |
| SA*sex | 0.87 | 0.75 | 1.00 | -0.14 | 0.07 | -1.93 | 1.000 |
| PA*sex | 1.06 | 0.96 | 1.19 | 0.06 | 0.06 | 1.14 | 1.000 |
| EA*sex | 0.93 | 0.84 | 1.03 | -0.07 | 0.05 | -1.40 | 1.000 |
| EN*sex | 1.06 | 0.95 | 1.18 | 0.06 | 0.05 | 1.07 | 1.000 |
| PN*sex | 1.12 | 0.99 | 1.26 | 0.11 | 0.06 | 1.81 | 1.000 |

*Note.* Sensitivity analysis with relative income position as additional covariate. OR = Odds Ratio. CTS = Childhood Trauma Screener. SA = Sexual Abuse. PA = Physical Abuse. EA = Emotional Abuse. EN = Emotional Neglect. PN = Physical Neglect. MINI = Mini International Neuropsychiatric Interview. PHQ-9 = Patient Health Questionnaire. Odds Ratios with 95% confidence intervals are presented for categorical measures. As reference group, participants who reported no/low trauma were selected. P_adj_ = adjusted *p*-values. ** *p_adj_* < .001 * *p_adj_* < .05. Adjusted *p*-values exceeding 1 are reported as *p* = 1.000.

### Table S11. *Sensitivity Analysis of the Multiple Binary Logistic Regression Analysis with Depression Measures as Outcome Variables in the L2 Subset*

| Variables | *OR* | *OR 95%*  *CI lower* | *OR 95%*  *CI upper* | *Estimate* | *Std. Error* | *z-Value* | *P_adj_* |
| --- | --- | --- | --- | --- | --- | --- | --- |
| **Physician’s diagnosis** |  |  |  |  |  |  |  |
| CTS | 2.42 | 2.28 | 2.57 | 0.89 | 0.03 | 29.37 | < .001** |
| CTS SA | 3.29 | 3.00 | 3.61 | 1.19 | 0.05 | 25.38 | < .001** |
| CTS PA | 2.40 | 2.21 | 2.61 | 0.88 | 0.04 | 20.36 | < .001** |
| CTS EA | 3.37 | 3.11 | 3.65 | 1.21 | 0.04 | 29.32 | < .001** |
| CTS EN | 3.21 | 2.95 | 3.49 | 1.17 | 0.04 | 27.14 | < .001** |
| CTS PN | 1.10 | 1.00 | 1.20 | 0.09 | 0.05 | 1.95 | .919 |
| **PHQ-9** |  |  |  |  |  |  |  |
| CTS | 3.02 | 2.79 | 3.26 | 1.10 | 0.04 | 28.33 | < .001** |
| CTS SA | 3.12 | 2.78 | 3.50 | 1.14 | 0.06 | 19.62 | < .001** |
| CTS PA | 2.73 | 2.46 | 3.03 | 1.01 | 0.05 | 18.85 | < .001** |
| CTS EA | 3.97 | 3.60 | 4.36 | 1.38 | 0.05 | 28.40 | < .001** |
| CTS EN | 3.75 | 3.39 | 4.16 | 1.32 | 0.05 | 25.21 | < .001** |
| CTS PN | 1.44 | 1.28 | 1.61 | 0.36 | 0.06 | 6.12 | < .001** |

*Note.* OR = Odds Ratio. CTS = Childhood Trauma Screener. SA = Sexual Abuse. PA = Physical Abuse. EA = Emotional Abuse. EN = Emotional Neglect. PN = Physical Neglect. MINI = Mini International Neuropsychiatric Interview. PHQ-9 = Patient Health Questionnaire. Odds Ratios with 95% confidence intervals are presented for categorical measures. As reference group, participants who reported no/low trauma were selected. P_adj_ = adjusted *p*-values. ** *p_adj_* < .001 * *p_adj_* < .05. Adjusted *p*-values exceeding 1 are reported as *p* = 1.000.

### Table S12. *Sensitivity Analysis of the Sex-Stratified Binary Logistic Regression Analysis with Depression Measures as Outcome Variables for Females in the L2 Subset*

| Variables | *OR* | *OR 95%*  *CI lower* | *OR 95%*  *CI upper* | *Estimate* | *Std. Error* | *z-Value* | *P_adj_* |
| --- | --- | --- | --- | --- | --- | --- | --- |
| **Physician’s diagnosis** |  |  |  |  |  |  |  |
| CTS | 2.54 | 2.36 | 2.75 | 0.93 | 0.04 | 23.73 | < .001** |
| CTS SA | 2.68 | 2.41 | 2.98 | 0.99 | 0.05 | 18.35 | < .001** |
| CTS PA | 2.63 | 2.35 | 2.95 | 0.97 | 0.06 | 16.62 | < .001** |
| CTS EA | 3.12 | 2.82 | 3.46 | 1.14 | 0.05 | 21.67 | < .001** |
| CTS EN | 3.30 | 2.96 | 3.68 | 1.19 | 0.06 | 21.62 | < .001** |
| CTS PN | 1.24 | 1.10 | 1.39 | 0.21 | 0.06 | 3.52 | .015* |
| **PHQ-9** |  |  |  |  |  |  |  |
| CTS | 2.97 | 2.69 | 3.29 | 1.09 | 0.05 | 21.17 | < .001** |
| CTS SA | 2.70 | 2.37 | 3.07 | 0.99 | 0.07 | 14.88 | < .001** |
| CTS PA | 2.70 | 2.34 | 3.10 | 0.99 | 0.07 | 13.80 | < .001** |
| CTS EA | 3.75 | 3.32 | 4.23 | 1.32 | 0.06 | 21.42 | < .001** |
| CTS EN | 3.56 | 3.11 | 4.06 | 1.27 | 0.07 | 18.69 | < .001** |
| CTS PN | 1.52 | 1.30 | 1.76 | 0.42 | 0.08 | 5.43 | < .001** |

*Note.* OR = Odds Ratio. CTS = Childhood Trauma Screener. SA = Sexual Abuse. PA = Physical Abuse. EA = Emotional Abuse. EN = Emotional Neglect. PN = Physical Neglect. MINI = Mini International Neuropsychiatric Interview. PHQ-9 = Patient Health Questionnaire. Odds Ratios with 95% confidence intervals are presented for categorical measures. As reference group, participants who reported no/low trauma were selected. P_adj_ = adjusted *p*-values. ** *p_adj_* < .001 * *p_adj_* < .05. Adjusted *p*-values exceeding 1 are reported as *p* = 1.000.

### Table S13. *Sensitivity Analysis of the Sex-Stratified Binary Logistic Regression Analysis with Depression Measures as Outcome Variables for Males in the L2 Subset*

| Variables | *OR* | *OR 95%*  *CI lower* | *OR 95%*  *CI upper* | *Estimate* | *Std. Error* | *z-Value* | *P_adj_* |
| --- | --- | --- | --- | --- | --- | --- | --- |
| **Physician’s diagnosis** |  |  |  |  |  |  |  |
| CTS | 2.11 | 1.92 | 2.31 | 0.75 | 0.05 | 15.54 | < .001** |
| CTS SA | 3.42 | 2.79 | 4.17 | 1.23 | 0.1 | 12.02 | < .001** |
| CTS PA | 2.29 | 2.01 | 2.60 | 0.83 | 0.07 | 12.57 | < .001** |
| CTS EA | 3.31 | 2.89 | 3.78 | 1.20 | 0.07 | 17.47 | < .001** |
| CTS EN | 2.85 | 2.48 | 3.27 | 1.05 | 0.07 | 14.93 | < .001** |
| CTS PN | 0.92 | 0.78 | 1.06 | -0.09 | 0.08 | -1.14 | 1.000 |
| **PHQ-9** |  |  |  |  |  |  |  |
| CTS | 2.92 | 2.60 | 3.29 | 1.07 | 0.06 | 17.82 | < .001** |
| CTS SA | 3.29 | 2.56 | 4.18 | 1.19 | 0.13 | 9.48 | < .001** |
| CTS PA | 2.88 | 2.45 | 3.36 | 1.06 | 0.08 | 13.15 | < .001** |
| CTS EA | 3.92 | 3.35 | 4.58 | 1.37 | 0.08 | 17.24 | < .001** |
| CTS EN | 3.82 | 3.24 | 4.49 | 1.34 | 0.08 | 16.12 | < .001** |
| CTS PN | 1.33 | 1.11 | 1.60 | 0.29 | 0.09 | 3.07 | .078 |

*Note.* OR = Odds Ratio. CTS = Childhood Trauma Screener. SA = Sexual Abuse. PA = Physical Abuse. EA = Emotional Abuse. EN = Emotional Neglect. PN = Physical Neglect. MINI = Mini International Neuropsychiatric Interview. PHQ-9 = Patient Health Questionnaire. Odds Ratios with 95% confidence intervals are presented for categorical measures. As reference group, participants who reported no/low trauma were selected. P_adj_ = adjusted *p*-values. ** *p_adj_* < .001 * *p_adj_* < .05. Adjusted *p*-values exceeding 1 are reported as *p* = 1.000.

### Table S14. *Sensitivity Analysis of the Multiple Binary Logistic Regression Analysis with Sex Interactions with Depression Measures as Outcome Variables in L2 Participants Only*

| Variables | *OR* | *OR 95%*  *CI lower* | *OR 95%*  *CI upper* | *Estimate* | *Std. Error* | *z-Value* | *P_adj_* |
| --- | --- | --- | --- | --- | --- | --- | --- |
| **Physician’s diagnosis** |  |  |  |  |  |  |  |
| CTS*sex | 1.21 | 1.08 | 1.37 | 0.19 | 0.06 | 3.17 | .027* |
| SA*sex | 0.78 | 0.62 | 0.98 | -0.25 | 0.12 | -2.15 | .575 |
| PA*sex | 1.16 | 0.98 | 1.38 | 0.15 | 0.09 | 1.70 | 1.000 |
| EA*sex | 0.92 | 0.78 | 1.09 | -0.08 | 0.09 | -0.92 | 1.000 |
| EN*sex | 1.16 | 0.97 | 1.38 | 0.15 | 0.09 | 1.64 | 1.000 |
| PN*sex | 1.36 | 1.12 | 1.65 | 0.31 | 0.10 | 3.14 | .030* |
| **PHQ-9** |  |  |  |  |  |  |  |
| CTS*sex | 1.06 | 0.91 | 1.24 | 0.06 | 0.08 | 0.78 | 1.000 |
| SA*sex | 0.83 | 0.63 | 1.11 | -0.18 | 0.14 | -1.28 | 1.000 |
| PA*sex | 0.99 | 0.80 | 1.22 | -0.01 | 0.11 | -0.12 | 1.000 |
| EA*sex | 0.96 | 0.79 | 1.17 | -0.04 | 0.10 | -0.37 | 1.000 |
| EN*sex | 0.97 | 0.79 | 1.20 | -0.03 | 0.11 | -0.24 | 1.000 |
| PN*sex | 1.19 | 0.94 | 1.51 | 0.17 | 0.12 | 1.46 | 1.000 |

*Note.* OR = Odds Ratio. CTS = Childhood Trauma Screener. SA = Sexual Abuse. PA = Physical Abuse. EA = Emotional Abuse. EN = Emotional Neglect. PN = Physical Neglect. MINI = Mini International Neuropsychiatric Interview. PHQ-9 = Patient Health Questionnaire. Odds Ratios with 95% confidence intervals are presented for categorical measures. As reference group, participants who reported no/low trauma were selected. P_adj_ = adjusted *p*-values. ** *p_adj_* < .001 * *p_adj_* < .05. Adjusted *p*-values exceeding 1 are reported as *p* = 1.000.

### Table S15. *Sensitivity Analysis of the Multiple Binary Logistic Regression Analysis with Depression Measures as Outcome Variables in Females – Different CTS Subtype Levels*

| Variables | *OR* | *OR 95%*  *CI lower* | *OR 95%*  *CI upper* | *Estimate* | *Std. Error* | *z-Value* | *P_adj_* |
| --- | --- | --- | --- | --- | --- | --- | --- |
| **Physician’s diagnosis** |  |  |  |  |  |  |  |
| SA  Rarely | 2.27 | 2.11 | 2.44 | 0.82 | 0.04 | 22.29 | < .001** |
| SA  Sometimes | 2.75 | 2.53 | 2.98 | 1.01 | 0.04 | 24.01 | < .001** |
| SA  Often | 3.57 | 3.01 | 4.24 | 1.27 | 0.09 | 14.61 | < .001** |
| SA  Very often | 4.00 | 3.33 | 4.80 | 1.39 | 0.09 | 14.78 | < .001** |
| PA  Rarely | 1.70 | 1.61 | 1.81 | 0.53 | 0.03 | 17.68 | < .001** |
| PA  Sometimes | 2.50 | 2.33 | 2.68 | 0.92 | 0.04 | 25.05 | < .001** |
| PA  Often | 3.44 | 3.09 | 3.84 | 1.24 | 0.06 | 22.12 | < .001** |
| PA  Very often | 4.35 | 3.79 | 4.99 | 1.47 | 0.07 | 20.97 | < .001** |
| EA  Rarely | 2.15 | 2.02 | 2.29 | 0.77 | 0.03 | 24.82 | < .001** |
| EA  Sometimes | 2.88 | 2.68 | 3.08 | 1.06 | 0.04 | 29.96 | < .001** |
| EA  Often | 4.78 | 4.38 | 5.22 | 1.57 | 0.04 | 35.17 | < .001** |
| EA  Very often | 4.36 | 3.91 | 4.85 | 1.47 | 0.05 | 26.83 | < .001** |
| EN  Rarely | 1.79 | 1.71 | 1.87 | 0.58 | 0.02 | 25.10 | < .001** |
| EN  Sometimes | 3.70 | 3.48 | 3.92 | 1.31 | 0.03 | 43.47 | < .001** |
| EN  Often | 4.86 | 4.55 | 5.18 | 1.58 | 0.03 | 47.51 | < .001** |
| EN  Very often | 5.76 | 5.17 | 6.41 | 1.75 | 0.05 | 31.88 | < .001** |
| PN  Rarely | 1.62 | 1.55 | 1.70 | 0.48 | 0.02 | 19.95 | < .001** |
| PN  Sometimes | 1.63 | 1.54 | 1.72 | 0.49 | 0.03 | 17.00 | < .001** |
| PN  Often | 1.90 | 1.76 | 2.06 | 0.64 | 0.04 | 15.85 | < .001** |
| PN  Very often | 1.45 | 1.34 | 1.57 | 0.37 | 0.04 | 9.00 | < .001** |
| **MINI** |  |  |  |  |  |  |  |
| SA  Rarely | 2.09 | 1.81 | 2.42 | 0.74 | 0.07 | 10.06 | < .001** |
| SA  Sometimes | 3.10 | 2.64 | 3.64 | 1.13 | 0.08 | 13.80 | < .001** |
| SA  Often | 3.29 | 2.33 | 4.65 | 1.19 | 0.18 | 6.78 | < .001** |
| SA  Very often | 2.69 | 1.78 | 4.04 | 0.99 | 0.21 | 4.74 | < .001** |
| PA  Rarely | 1.97 | 1.76 | 2.21 | 0.68 | 0.06 | 11.89 | < .001** |
| PA  Sometimes | 2.52 | 2.19 | 2.89 | 0.92 | 0.07 | 12.90 | < .001** |
| PA  Often | 2.58 | 2.07 | 3.22 | 0.95 | 0.11 | 8.40 | < .001** |
| PA  Very often | 4.35 | 3.29 | 5.77 | 1.47 | 0.14 | 10.24 | < .001** |
| EA  Rarely | 2.26 | 2.01 | 2.53 | 0.81 | 0.06 | 13.98 | < .001** |
| EA  Sometimes | 2.92 | 2.55 | 3.35 | 1.07 | 0.07 | 15.54 | < .001** |
| EA  Often | 3.97 | 3.33 | 4.73 | 1.38 | 0.09 | 15.32 | < .001** |
| EA  Very often | 3.85 | 3.09 | 4.79 | 1.35 | 0.11 | 12.00 | < .001** |
| EN  Rarely | 1.74 | 1.60 | 1.90 | 0.56 | 0.04 | 12.85 | < .001** |
| EN  Sometimes | 3.06 | 2.72 | 3.44 | 1.12 | 0.06 | 18.78 | < .001** |
| EN  Often | 4.26 | 3.75 | 4.85 | 1.45 | 0.07 | 22.05 | < .001** |
| EN  Very often | 4.39 | 3.55 | 5.44 | 1.48 | 0.11 | 13.56 | < .001** |
| PN  Rarely | 1.50 | 1.37 | 1.64 | 0.40 | 0.05 | 8.69 | < .001** |
| PN  Sometimes | 1.53 | 1.37 | 1.71 | 0.43 | 0.06 | 7.63 | < .001** |
| PN  Often | 1.54 | 1.31 | 1.82 | 0.43 | 0.08 | 5.19 | < .001** |
| PN  Very often | 1.34 | 1.14 | 1.57 | 0.29 | 0.08 | 3.58 | .012* |
| **PHQ-9** |  |  |  |  |  |  |  |
| SA  Rarely | 2.21 | 2.02 | 2.42 | 0.79 | 0.05 | 17.01 | < .001** |
| SA  Sometimes | 2.92 | 2.64 | 3.22 | 1.07 | 0.05 | 21.03 | < .001** |
| SA  Often | 3.68 | 3.03 | 4.48 | 1.30 | 0.10 | 13.06 | < .001** |
| SA  Very often | 5.15 | 4.22 | 6.28 | 1.64 | 0.10 | 16.16 | < .001** |
| PA  Rarely | 1.77 | 1.63 | 1.91 | 0.57 | 0.04 | 14.37 | < .001** |
| PA  Sometimes | 2.65 | 2.43 | 2.90 | 0.98 | 0.05 | 21.40 | < .001** |
| PA  Often | 3.36 | 2.95 | 3.83 | 1.21 | 0.07 | 18.11 | < .001** |
| PA  Very often | 5.15 | 4.43 | 5.98 | 1.64 | 0.08 | 21.30 | < .001** |
| EA  Rarely | 2.43 | 2.25 | 2.62 | 0.89 | 0.04 | 22.92 | < .001** |
| EA  Sometimes | 3.34 | 3.08 | 3.63 | 1.21 | 0.04 | 28.48 | < .001** |
| EA  Often | 4.92 | 4.46 | 5.44 | 1.59 | 0.05 | 31.29 | < .001** |
| EA  Very often | 5.73 | 5.09 | 6.44 | 1.74 | 0.06 | 29.03 | < .001** |
| EN  Rarely | 1.81 | 1.71 | 1.93 | 0.60 | 0.03 | 18.78 | < .001** |
| EN  Sometimes | 3.52 | 3.25 | 3.80 | 1.26 | 0.04 | 31.73 | < .001** |
| EN  Often | 5.27 | 4.86 | 5.72 | 1.66 | 0.04 | 39.84 | < .001** |
| EN  Very often | 7.60 | 6.73 | 8.59 | 2.03 | 0.06 | 32.54 | < .001** |
| PN  Rarely | 1.69 | 1.59 | 1.80 | 0.52 | 0.03 | 16.34 | < .001** |
| PN  Sometimes | 1.82 | 1.69 | 1.96 | 0.60 | 0.04 | 15.66 | < .001** |
| PN  Often | 2.21 | 1.99 | 2.45 | 0.79 | 0.05 | 15.13 | < .001** |
| PN  Very often | 1.56 | 1.40 | 1.74 | 0.44 | 0.06 | 7.98 | < .001** |

*Note.* OR = Odds Ratio. CTS = Childhood Trauma Screener. SA = Sexual Abuse. PA = Physical Abuse. EA = Emotional Abuse. EN = Emotional Neglect. PN = Physical Neglect. MINI = Mini International Neuropsychiatric Interview. PHQ-9 = Patient Health Questionnaire. Odds Ratios with 95% confidence intervals are presented for categorical measures. As reference group, participants who reported no/low trauma were selected. P_adj_ = adjusted *p*-values. ** *p_adj_* < .001 * *p_adj_* < .05. Adjusted *p*-values exceeding 1 are reported as *p* = 1.000.

### Table S16. *Sensitivity Analysis of the Multiple Binary Logistic Regression Analysis with Depression Measures as Outcome Variables in Males – Different CTS Subtype Levels*

| Variables | *OR* | *OR 95%*  *CI lower* | *OR 95%*  *CI upper* | *Estimate* | *Std. Error* | *z-Value* | *P_adj_* |
| --- | --- | --- | --- | --- | --- | --- | --- |
| **Physician’s diagnosis** |  |  |  |  |  |  |  |
| SA  Rarely | 2.80 | 2.38 | 3.30 | 1.03 | 0.08 | 12.40 | < .001** |
| SA  Sometimes | 3.92 | 3.16 | 4.86 | 1.37 | 0.11 | 12.38 | < .001** |
| SA  Often | 4.25 | 2.72 | 6.64 | 1.45 | 0.23 | 6.35 | < .001** |
| SA  Very often | 4.66 | 3.00 | 7.26 | 1.54 | 0.23 | 6.82 | < .001** |
| PA  Rarely | 1.81 | 1.67 | 1.96 | 0.59 | 0.04 | 14.34 | < .001** |
| PA  Sometimes | 2.61 | 2.36 | 2.89 | 0.96 | 0.05 | 18.66 | < .001** |
| PA  Often | 4.39 | 3.80 | 5.08 | 1.48 | 0.07 | 19.96 | < .001** |
| PA  Very often | 4.45 | 3.79 | 5.46 | 1.49 | 0.10 | 14.24 | < .001** |
| EA  Rarely | 2.48 | 2.28 | 2.71 | 0.91 | 0.04 | 20.28 | < .001** |
| EA  Sometimes | 3.77 | 3.39 | 4.19 | 1.33 | 0.05 | 24.58 | < .001** |
| EA  Often | 5.98 | 5.22 | 6.84 | 1.79 | 0.07 | 25.86 | < .001** |
| EA  Very often | 5.22 | 4.34 | 6.27 | 1.65 | 0.09 | 17.67 | < .001** |
| EN  Rarely | 1.52 | 1.41 | 1.63 | 0.42 | 0.04 | 11.24 | < .001** |
| EN  Sometimes | 3.33 | 3.04 | 3.65 | 1.20 | 0.05 | 25.54 | < .001** |
| EN  Often | 5.05 | 4.55 | 5.60 | 1.62 | 0.05 | 30.74 | < .001** |
| EN  Very often | 5.83 | 4.97 | 6.83 | 1.76 | 0.08 | 21.70 | < .001** |
| PN  Rarely | 1.54 | 1.43 | 1.66 | 0.43 | 0.04 | 11.52 | < .001** |
| PN  Sometimes | 1.47 | 1.34 | 1.61 | 0.39 | 0.05 | 8.31 | < .001** |
| PN  Often | 1.85 | 1.62 | 2.10 | 0.61 | 0.07 | 9.34 | < .001** |
| PN  Very often | 1.40 | 1.23 | 1.60 | 0.34 | 0.07 | 5.04 | < .001** |
| **MINI** |  |  |  |  |  |  |  |
| SA  Rarely | 2.56 | 1.99 | 3.29 | 0.94 | 0.13 | 7.31 | < .001** |
| SA  Sometimes | 3.31 | 2.35 | 4.66 | 1.20 | 0.17 | 6.87 | < .001** |
| SA  Often | 3.55 | 1.57 | 8.02 | 1.27 | 0.42 | 3.05 | .083 |
| SA  Very often | 2.12 | 0.79 | 5.75 | 0.75 | 0.51 | 1.48 | 1.000 |
| PA  Rarely | 1.54 | 1.38 | 1.71 | 0.43 | 0.05 | 7.82 | < .001** |
| PA  Sometimes | 2.10 | 1.82 | 2.43 | 0.74 | 0.07 | 10.09 | < .001** |
| PA  Often | 3.40 | 2.70 | 4.30 | 1.23 | 0.12 | 10.32 | < .001** |
| PA  Very often | 2.90 | 2.03 | 4.13 | 1.06 | 0.18 | 5.85 | < .001** |
| EA  Rarely | 2.16 | 1.92 | 2.44 | 0.77 | 0.06 | 12.43 | < .001** |
| EA  Sometimes | 3.42 | 2.92 | 4.01 | 1.23 | 0.08 | 15.16 | < .001** |
| EA  Often | 4.72 | 3.79 | 5.86 | 1.55 | 0.11 | 13.97 | < .001** |
| EA  Very often | 3.07 | 2.20 | 4.28 | 1.12 | 0.17 | 6.58 | < .001** |
| EN  Rarely | 1.58 | 1.44 | 1.73 | 0.45 | 0.05 | 9.80 | < .001** |
| EN  Sometimes | 3.03 | 2.68 | 3.43 | 1.11 | 0.06 | 17.46 | < .001** |
| EN  Often | 3.84 | 3.30 | 4.48 | 1.35 | 0.08 | 17.29 | < .001** |
| EN  Very often | 3.55 | 2.70 | 4.67 | 1.27 | 0.14 | 9.07 | < .001** |
| PN  Rarely | 1.38 | 1.25 | 1.52 | 0.32 | 0.05 | 6.49 | < .001** |
| PN  Sometimes | 1.33 | 1.18 | 1.49 | 0.28 | 0.06 | 4.68 | < .001** |
| PN  Often | 1.30 | 1.08 | 1.57 | 0.26 | 0.10 | 2.76 | .209 |
| PN  Very often | 1.07 | 0.89 | 1.29 | 0.07 | 0.10 | 0.69 | 1.000 |
| **PHQ-9** |  |  |  |  |  |  |  |
| SA  Rarely | 2.80 | 2.38 | 3.30 | 1.03 | 0.08 | 12.40 | < .001** |
| SA  Sometimes | 3.92 | 3.16 | 4.86 | 1.37 | 0.11 | 12.38 | < .001** |
| SA  Often | 4.25 | 2.72 | 6.64 | 1.45 | 0.23 | 6.35 | < .001** |
| SA  Very often | 4.66 | 3.00 | 7.26 | 1.54 | 0.23 | 6.82 | < .001** |
| PA  Rarely | 1.81 | 1.67 | 1.96 | 0.59 | 0.04 | 14.34 | < .001** |
| PA  Sometimes | 2.61 | 2.36 | 2.89 | 0.96 | 0.05 | 18.66 | < .001** |
| PA  Often | 4.39 | 3.80 | 5.08 | 1.48 | 0.07 | 19.96 | < .001** |
| PA  Very often | 4.45 | 3.62 | 5.46 | 1.49 | 0.10 | 14.24 | < .001** |
| EA  Rarely | 2.48 | 2.28 | 2.71 | 0.91 | 0.04 | 20.28 | < .001** |
| EA  Sometimes | 3.77 | 3.39 | 4.19 | 1.33 | 0.05 | 24.58 | < .001** |
| EA  Often | 5.98 | 5.22 | 6.84 | 1.79 | 0.07 | 25.86 | < .001** |
| EA  Very often | 5.22 | 4.34 | 6.27 | 1.65 | 0.09 | 17.67 | < .001** |
| EN  Rarely | 1.52 | 1.41 | 1.63 | 0.42 | 0.04 | 11.24 | < .001** |
| EN  Sometimes | 3.33 | 3.04 | 3.65 | 1.20 | 0.05 | 25.54 | < .001** |
| EN  Often | 5.05 | 4.55 | 5.60 | 1.62 | 0.05 | 30.74 | < .001** |
| EN  Very often | 5.83 | 4.97 | 6.83 | 1.76 | 0.08 | 21.70 | < .001** |
| PN  Rarely | 1.54 | 1.43 | 1.66 | 0.43 | 0.04 | 11.52 | < .001** |
| PN  Sometimes | 1.47 | 1.34 | 1.61 | 0.39 | 0.05 | 8.31 | < .001** |
| PN  Often | 1.85 | 1.62 | 2.10 | 0.61 | 0.07 | 9.34 | < .001** |
| PN  Very often | 1.40 | 1.23 | 1.60 | 0.34 | 0.07 | 5.04 | < .001** |

*Note.* OR = Odds Ratio. CTS = Childhood Trauma Screener. SA = Sexual Abuse. PA = Physical Abuse. EA = Emotional Abuse. EN = Emotional Neglect. PN = Physical Neglect. MINI = Mini International Neuropsychiatric Interview. PHQ-9 = Patient Health Questionnaire. Odds Ratios with 95% confidence intervals are presented for categorical measures. As reference group, participants who reported no/low trauma were selected. P_adj_ = adjusted *p*-values. ** *p_adj_* < .001 * *p_adj_* < .05. Adjusted *p*-values exceeding 1 are reported as *p* = 1.000.

### Table S17. *Sensitivity Analysis of the Multiple Binary Logistic Regression Analysis with Sex Interactions with Depression Measures as Outcome Variables – Different CTS Subtype Levels*

| Variables | *OR* | *OR 95%*  *CI lower* | *OR 95%*  *CI upper* | *Estimate* | *Std. Error* | *z-Value* | *P_adj_* |
| --- | --- | --- | --- | --- | --- | --- | --- |
| **Physician’s diagnosis** |  |  |  |  |  |  |  |
| SA*sex  Rarely | 0.77 | 0.66 | 0.90 | -0.26 | 0.08 | -3.39 | .013* |
| SA*sex  Sometimes | 0.74 | 0.60 | 0.90 | -0.30 | 0.10 | -2.96 | .056 |
| SA*sex  Often | 0.77 | 0.50 | 1.17 | -0.26 | 0.22 | -1.23 | 1.000 |
| SA*sex  Very often | 1.23 | 0.79 | 1.91 | 0.21 | 0.23 | 0.91 | 1.000 |
| PA*sex  Rarely | 1.16 | 1.07 | 1.27 | 0.15 | 0.04 | 3.41 | .012* |
| PA*sex  Sometimes | 1.14 | 1.03 | 1.27 | 0.13 | 0.05 | 2.46 | .248 |
| PA*sex  Often | 1.00 | 0.84 | 1.18 | 0.00 | 0.08 | -0.04 | 1.000 |
| PA*sex  Very often | 1.22 | 0.97 | 1.53 | 0.20 | 0.11 | 1.73 | 1.000 |
| EA*sex  Rarely | 1.02 | 0.93 | 1.12 | 0.02 | 0.05 | 0.43 | 1.000 |
| EA*sex  Sometimes | 0.88 | 0.79 | 0.99 | -0.12 | 0.06 | -2.14 | .581 |
| EA*sex  Often | 0.97 | 0.84 | 1.13 | -0.03 | 0.08 | -0.33 | 1.000 |
| EA*sex  Very often | 1.21 | 0.99 | 1.47 | 0.19 | 0.10 | 1.84 | 1.000 |
| EN*sex  Rarely | 1.21 | 1.13 | 1.30 | 0.19 | 0.04 | 5.30 | < .001** |
| EN*sex  Sometimes | 1.24 | 1.13 | 1.36 | 0.22 | 0.05 | 4.58 | < .001** |
| EN*sex  Often | 1.09 | 0.98 | 1.21 | 0.08 | 0.05 | 1.59 | 1.000 |
| EN*sex  Very often | 1.37 | 1.15 | 1.64 | 0.32 | 0.09 | 3.54 | .007 |
| PN*sex  Rarely | 1.13 | 1.05 | 1.22 | 0.13 | 0.04 | 3.31 | .017* |
| PN*sex  Sometimes | 1.19 | 1.09 | 1.30 | 0.18 | 0.04 | 3.95 | .001** |
| PN*sex  Often | 1.40 | 1.23 | 1.59 | 0.34 | 0.07 | 5.06 | < .001** |
| PN*sex  Very often | 1.19 | 1.05 | 1.36 | 0.18 | 0.07 | 2.68 | .132 |
| **MINI** |  |  |  |  |  |  |  |
| SA*sex  Rarely | 0.82 | 0.61 | 1.09 | -0.20 | 0.15 | -1.38 | 1.000 |
| SA*sex  Sometimes | 0.93 | 0.64 | 1.35 | -0.07 | 0.19 | -0.38 | 1.000 |
| SA*sex  Often | 0.87 | 0.36 | 2.10 | -0.14 | 0.45 | -0.31 | 1.000 |
| SA*sex  Very often | 1.24 | 0.42 | 3.64 | 0.22 | 0.55 | 0.40 | 1.000 |
| PA*sex  Rarely | 1.31 | 1.12 | 1.53 | 0.27 | 0.08 | 3.40 | .012* |
| PA*sex  Sometimes | 1.21 | 0.99 | 1.48 | 0.19 | 0.10 | 1.87 | 1.000 |
| PA*sex  Often | 0.75 | 0.55 | 1.04 | -0.28 | 0.16 | -1.74 | 1.000 |
| PA*sex  Very often | 1.42 | 0.90 | 2.23 | 0.35 | 0.23 | 1.51 | 1.000 |
| EA*sex  Rarely | 1.03 | 0.87 | 1.21 | 0.03 | 0.09 | 0.32 | 1.000 |
| EA*sex  Sometimes | 0.83 | 0.67 | 1.02 | -0.19 | 0.11 | -1.78 | 1.000 |
| EA*sex  Often | 0.81 | 0.61 | 1.07 | -0.21 | 0.14 | -1.46 | 1.000 |
| EA*sex  Very often | 1.17 | 0.79 | 1.75 | 0.16 | 0.20 | 0.79 | 1.000 |
| EN*sex  Rarely | 1.12 | 0.99 | 1.27 | 0.11 | 0.06 | 1.78 | 1.000 |
| EN*sex  Sometimes | 1.02 | 0.86 | 1.21 | 0.02 | 0.09 | 0.20 | 1.000 |
| EN*sex  Often | 1.11 | 0.91 | 1.36 | 0.11 | 0.10 | 1.07 | 1.000 |
| EN*sex  Very often | 1.17 | 0.83 | 1.65 | 0.16 | 0.18 | 0.90 | 1.000 |
| PN*sex  Rarely | 1.08 | 0.95 | 1.24 | 0.08 | 0.07 | 1.20 | 1.000 |
| PN*sex  Sometimes | 1.17 | 1.00 | 1.38 | 0.16 | 0.08 | 2.00 | .810 |
| PN*sex  Often | 1.20 | 0.94 | 1.53 | 0.18 | 0.12 | 1.43 | 1.000 |
| PN*sex  Very often | 1.24 | 0.97 | 1.58 | 0.21 | 0.12 | 1.72 | 1.000 |
| **PHQ-9** |  |  |  |  |  |  |  |
| SA*sex  Rarely | 0.79 | 0.66 | 0.95 | -0.23 | 0.10 | -2.46 | .247 |
| SA*sex  Sometimes | 0.74 | 0.59 | 0.94 | -0.30 | 0.12 | -2.45 | .259 |
| SA*sex  Often | 0.85 | 0.52 | 1.38 | -0.16 | 0.25 | -0.66 | 1.000 |
| SA*sex  Very often | 1.09 | 0.67 | 1.76 | 0.08 | 0.25 | 0.34 | 1.000 |
| PA*sex  Rarely | 0.99 | 0.89 | 1.11 | -0.01 | 0.06 | -0.13 | 1.000 |
| PA*sex  Sometimes | 1.03 | 0.90 | 1.18 | 0.03 | 0.07 | 0.48 | 1.000 |
| PA*sex  Often | 0.77 | 0.64 | 0.94 | -0.26 | 0.10 | -2.61 | .165 |
| PA*sex  Very often | 1.16 | 0.90 | 1.49 | 0.15 | 0.13 | 1.14 | 1.000 |
| EA*sex  Rarely | 0.97 | 0.87 | 1.09 | -0.03 | 0.06 | -0.48 | 1.000 |
| EA*sex  Sometimes | 0.88 | 0.77 | 1.01 | -0.13 | 0.07 | -1.88 | 1.000 |
| EA*sex  Often | 0.82 | 0.69 | 0.97 | -0.20 | 0.09 | -2.37 | .324 |
| EA*sex  Very often | 1.08 | 0.87 | 1.35 | 0.08 | 0.11 | 0.73 | 1.000 |
| EN*sex  Rarely | 1.20 | 1.09 | 1.32 | 0.18 | 0.05 | 3.70 | .004* |
| EN*sex  Sometimes | 1.06 | 0.94 | 1.19 | 0.06 | 0.06 | 0.93 | 1.000 |
| EN*sex  Often | 1.05 | 0.92 | 1.19 | 0.05 | 0.07 | 0.69 | 1.000 |
| EN*sex  Very often | 1.29 | 1.06 | 1.57 | 0.25 | 0.10 | 2.51 | 0.218 |
| PN*sex  Rarely | 1.09 | 0.99 | 1.20 | 0.08 | 0.05 | 1.72 | 1.000 |
| PN*sex  Sometimes | 1.23 | 1.10 | 1.38 | 0.21 | 0.06 | 3.53 | .008* |
| PN*sex  Often | 1.19 | 1.01 | 1.39 | 0.17 | 0.08 | 2.07 | .685 |
| PN*sex  Very often | 1.09 | 0.92 | 1.29 | 0.09 | 0.09 | 1.04 | 1.0000 |

*Note.* OR = Odds Ratio. CTS = Childhood Trauma Screener. SA = Sexual Abuse. PA = Physical Abuse. EA = Emotional Abuse. EN = Emotional Neglect. PN = Physical Neglect. MINI = Mini International Neuropsychiatric Interview. PHQ-9 = Patient Health Questionnaire. Odds Ratios with 95% confidence intervals are presented for categorical measures. As reference group, participants who reported no/low trauma were selected. P_adj_ = adjusted *p*-values. ** *p_adj_* < .001 * *p_adj_* < .05. Adjusted *p*-values exceeding 1 are reported as *p* = 1.000.

### Table S18. *Results of Population Attributable Fractions and Risk Ratios of Depression Cases Attributable to Childhood Maltreatment for the Total Sample*

|  | RR | | | C | PAF | |  | Indiv. weighted PAF | |
| --- | --- | --- | --- | --- | --- | --- | --- | --- | --- |
|  |  | | 95 % CI |  |  | 95 % CI |  |  | 95 % CI |
| **Physician’s diagnosis of depression** |  |  | |  |  |  |  |  |  |
| Sexual Abuse | 2.37 | | 2.30–2.45 | 0.14 | 7.6% | 7.2–8.0 |  | 5.8% | 5.5–6.1 |
| Physical Abuse | 2.02 | | 1.96–2.09 | 0.26 | 7.6% | 7.1–8.0 |  | 5.0% | 4.7–5.2 |
| Emotional Abuse | 2.57 | | 2.50–2.64 | 0.25 | 11.2% | 10.7–11.7 |  | 7.5% | 7.2–7.8 |
| Emotional Neglect | 2.48 | | 2.41–2.56 | 0.26 | 9.9% | 9.4–10.3 |  | 6.5% | 6.2–6.8 |
| Physical Neglect | 1.19 | | 1.14–1.23 | 0.08 | 1.8% | 1.4–2.2 |  | 1.5% | 1.1–1.8 |
| Overall PAF |  | |  |  |  |  |  | 26.2% | 25.4–27.1 |
| **MINI** |  | |  |  |  |  |  |  |  |
| Sexual Abuse | 2.19 | | 2.06–2.33 | 0.14 | 5.9% | 5.3–6.6 |  | 4.7% | 4.2–5.2 |
| Physical Abuse | 1.91 | | 1.80–2.02 | 0.27 | 6.3% | 5.6–7.1 |  | 4.2% | 3.7–4.7 |
| Emotional Abuse | 2.42 | | 2.30–2.55 | 0.26 | 9.4% | 8.6–10.2 |  | 6.4% | 5.8–6.9 |
| Emotional Neglect | 2.23 | | 2.11–2.37 | 0.26 | 7.6% | 6.9–8.4 |  | 5.1% | 4.7–5.7 |
| Physical Neglect | 1.10 | | 1.03–1.18 | 0.07 | 0.9% | 0.3–1.6 |  | 0.8% | 0.2–1.4 |
| Overall PAF |  | |  |  |  |  |  | 21.2% | 19.8–22.7 |
| **PHQ-9** |  | |  |  |  |  |  |  |  |
| Sexual Abuse | 2.58 | | 2.47–2.69 | 0.14 | 8.6% | 8.0–9.2 |  | 6.4% | 6.0–6.8 |
| Physical Abuse | 2.42 | | 2.32–2.53 | 0.26 | 10.2% | 9.5–10.9 |  | 6.5% | 6.1–6.9 |
| Emotional Abuse | 3.17 | | 3.05–3.28 | 0.25 | 14.8% | 14.1–15.6 |  | 9.6% | 9.1–10.0 |
| Emotional Neglect | 3.06 | | 2.94–3.18 | 0.26 | 13.8% | 12.4–13.9 |  | 8.4% | 8.0–8.8 |
| Physical Neglect | 1.35 | | 1.28–1.42 | 0.08 | 3.3% | 2.7–3.9 |  | 2.6% | 2.1–3.1 |
| Overall PAF |  | |  |  |  |  |  | 33.4% | 32.2–34.6 |

*Note.* RR = Risk Ratio. C = Communality. PAF = Population Attributable Fractions. MINI = Mini International Neuropsychiatric Interview. PHQ-9 = Patient Health Questionnaire. 95% confidence intervals are reported. Sum of individual PAF could differ from overall PAF estimates due to rounding*.*

### Table S19. *Results of Population Attributable Fractions and Risk Ratios of Depression Cases Attributable to Childhood Maltreatment for Females*

|  | RR | | | C | PAF | |  | Indiv. weighted PAF | |
| --- | --- | --- | --- | --- | --- | --- | --- | --- | --- |
|  |  | | 95 % CI |  |  | 95 % CI |  |  | 95 % CI |
| **Physician’s diagnosis of depression** |  |  | |  |  |  |  |  |  |
| Sexual Abuse | 2.02 | | 1.95–2.10 | 0.14 | 8.9% | 8.3–9.5 |  | 6.7% | 6.3–7.2 |
| Physical Abuse | 2.08 | | 2.00–2.16 | 0.25 | 7.7% | 7.2–8.2 |  | 5.1% | 4.7–5.4 |
| Emotional Abuse | 2.36 | | 2.29–2.45 | 0.26 | 12.0% | 11.3–12.6 |  | 7.9% | 7.5–8.2 |
| Emotional Neglect | 2.38 | | 2.30–2.47 | 0.25 | 10.3% | 9.7–11.0 |  | 6.8% | 6.4–7.2 |
| Physical Neglect | 1.27 | | 1.21–1.32 | 0.10 | 2.5% | 2.0–3.1 |  | 2.0% | 1.6–2.4 |
| Overall PAF |  | |  |  |  |  |  | 28.5% | 27.3–29.7 |
| **MINI** |  | |  |  |  |  |  |  |  |
| Sexual Abuse | 1.97 | | 1.84–2.12 | 0.13 | 8.0% | 6.9–9.1 |  | 6.3% | 5.4–7.1 |
| Physical Abuse | 1.96 | | 1.82–2.11 | 0.26 | 6.6% | 5.6–7.6 |  | 4.4% | 3.8–5.1 |
| Emotional Abuse | 2.24 | | 2.08–2.39 | 0.26 | 10.2% | 9.1–11.3 |  | 6.8% | 6.1–7.5 |
| Emotional Neglect | 2.21 | | 2.05–2.38 | 0.25 | 8.7% | 7.6–9.8 |  | 5.8% | 5.1–6.5 |
| Physical Neglect | 1.16 | | 1.06–1.27 | 0.09 | 1.5% | 0.5–2.5 |  | 1.2% | 0.5–2.0 |
| Overall PAF |  | |  |  |  |  |  | 24.5% | 22.4–26.6 |
| **PHQ-9** |  | |  |  |  |  |  |  |  |
| Sexual Abuse | 2.29 | | 2.18–2.42 | 0.14 | 11.0% | 10.1–11.9 |  | 8.0% | 7.4–8.6 |
| Physical Abuse | 2.41 | | 2.29–2.54 | 0.25 | 9.8% | 9.0–10.7 |  | 6.2% | 5.7–6.7 |
| Emotional Abuse | 2.93 | | 2.80–3.07 | 0.26 | 16.1% | 15.2–17.1 |  | 10.2% | 9.6–10.7 |
| Emotional Neglect | 2.96 | | 2.81–3.11 | 0.25 | 14.0% | 13.0–15.0 |  | 8.9% | 8.3–9.5 |
| Physical Neglect | 1.39 | | 1.30–1.48 | 0.10 | 3.7% | 2.8–4.5 |  | 2.8% | 2.2–3.4 |
| Overall PAF |  | |  |  |  |  |  | 36.1% | 34.5–37.7 |

*Note.* RR = Risk Ratio. C = Communality. PAF = Population Attributable Fractions. MINI = Mini International Neuropsychiatric Interview. PHQ-9 = Patient Health Questionnaire. 95% confidence intervals are presented. Sum of individual PAF could differ from overall PAF estimates due to rounding*.*

### Table S20. *Results of Population Attributable Fractions and Risk Ratios of Depression Cases Attributable to Childhood Maltreatment for Males*

|  | RR | | | C | PAF | |  | Indiv. weighted PAF | |
| --- | --- | --- | --- | --- | --- | --- | --- | --- | --- |
|  |  | | 95 % CI |  |  | 95 % CI |  |  | 95 % CI |
| **Physician’s diagnosis** |  |  | |  |  |  |  |  |  |
| Sexual Abuse | 2.51 | | 2.33–2.69 | 0.14 | 3.7% | 3.2–4.1 |  | 2.9% | 2.6–3.3 |
| Physical Abuse | 2.03 | | 1.93–2.14 | 0.28 | 7.9% | 7.1–8.6 |  | 5.2% | 4.8–5.7 |
| Emotional Abuse | 2.63 | | 2.51–2.77 | 0.26 | 9.2% | 8.5–9.9 |  | 6.2% | 5.8–6.7 |
| Emotional Neglect | 2.50 | | 2.38–2.63 | 0.26 | 8.8% | 8.1–9.6 |  | 6.0% | 5.5–6.5 |
| Physical Neglect | 1.07 | | 1.00–1.14 | 0.07 | 0.7% | 0.4–1.3 |  | 0.6% | 0.3–1.1 |
| Overall PAF |  | |  |  |  |  |  | 20.9% | 19.6–22.3 |
| **MINI** |  | |  |  |  |  |  |  |  |
| Sexual Abuse | 2.18 | | 1.90–2.47 | 0.12 | 2.5% | 1.9–3.2 |  | 2.1% | 1.6–2.6 |
| Physical Abuse | 1.88 | | 1.72–2.07 | 0.28 | 6.2% | 5.1–7.4 |  | 4.2% | 3.4–5.0 |
| Emotional Abuse | 2.52 | | 2.32–2.75 | 0.27 | 7.9% | 6.9–8.9 |  | 5.4% | 4.7–6.1 |
| Emotional Neglect | 2.14 | | 1.95–2.34 | 0.27 | 6.2% | 5.0–7.1 |  | 4.2% | 3.5–4.8 |
| Physical Neglect | 1.03 | | 0.92–1.14 | 0.06 | 0.2% | 0.1–1.3 |  | 0.2% | 0.1–1.1 |
| Overall PAF |  | |  |  |  |  |  | 16.0% | 14.1–18.0 |
| **PHQ-9** |  | |  |  |  |  |  |  |  |
| Sexual Abuse | 2.78 | | 2.51–3.05 | 0.14 | 4.3% | 3.7–4.9 |  | 3.3% | 2.8–3.7 |
| Physical Abuse | 2.48 | | 2.31–2.65 | 0.28 | 10.9% | 9.8–12.1 |  | 7.0% | 6.3–7.7 |
| Emotional Abuse | 3.32 | | 3.10–3.54 | 0.26 | 12.5% | 11.5–13.5 |  | 8.2% | 7.6–8.8 |
| Emotional Neglect | 3.06 | | 2.86–3.27 | 0.26 | 11.7% | 10.6–12.8 |  | 7.7% | 7.0–8.3 |
| Physical Neglect | 1.30 | | 1.19–1.41 | 0.07 | 2.8% | 1.8–3.9 |  | 2.3% | 1.5–3.2 |
| Overall PAF |  | |  |  |  |  |  | 28.4% | 26.7–30.2 |

*Note.* RR = Risk Ratios. C = Communality. PAF = Population Attributable Fractions. MINI = Mini International Neuropsychiatric Interview. PHQ-9 = Patient Health Questionnaire. 95% confidence intervals are presented. Sum of individual PAF could differ from overall PAF estimates due to rounding*.*

### Table S21. *Sex Differences in Population Attributable Fractions of Depression Cases Attributable to Childhood Maltreatment*

|  | Males | | | | | | | |  | Females | | | | |  |
| --- | --- | --- | --- | --- | --- | --- | --- | --- | --- | --- | --- | --- | --- | --- | --- |
|  | RR | | C | | indiv. weighted PAF | | | |  | RR | | C | indiv. weighted PAF | | *P_adj_* |
|  |  | 95 % CI |  | |  | | | 95 % CI |  |  | 95 % CI |  |  | 95 % CI |  |
| **PD** |  | | |  | |  |  |  |  |  |  |  |  |  |  |
| SA | 2.51 | 2.33–2.69 | 0.14 | | 2.9% | | | 2.6–3.3 |  | 2.02 | 1.95–2.10 | 0.14 | 6.7% | 6.3–7.2 | < .0072* |
| PA | 2.03 | 1.93–2.14 | 0.28 | | 5.2% | | | 4.8–5.7 |  | 2.08 | 2.00–2.16 | 0.25 | 5.1% | 4.7–5.4 | 1.000 |
| EA | 2.63 | 2.51–2.77 | 0.26 | | 6.2% | | | 5.8–6.7 |  | 2.36 | 2.29–2.45 | 0.26 | 7.9% | 7.5–8.2 | < .0072* |
| EN | 2.50 | 2.38–2.63 | 0.26 | | 6.0% | | | 5.5–6.5 |  | 2.38 | 2.30–2.47 | 0.25 | 6.8% | 6.4–7.2 | .0072* |
| PN | 1.07 | 1.00–1.14 | 0.07 | | 0.6% | | | 0.3–1.1 |  | 1.27 | 1.21–1.32 | 0.10 | 2.0% | 1.6–2.4 | < .0072* |
| PAF |  |  |  | | 20.9% | | | 19.6–22.3 |  |  |  |  | 28.5% | 27.3–29.7 | < .0072* |
| **MINI** |  |  |  | |  | | |  |  |  |  |  |  |  |  |
| SA | 2.18 | 1.90–2.47 | 0.12 | | 2.1% | | | 1.6–2.6 |  | 1.97 | 1.84–2.12 | 0.13 | 6.3% | 5.4–7.1 | < .0072* |
| PA | 1.88 | 1.72–2.07 | 0.28 | | 4.2% | | | 3.4–5.0 |  | 1.96 | 1.82–2.11 | 0.26 | 4.4% | 3.8–5.1 | 1.000 |
| EA | 2.52 | 2.32–2.75 | 0.27 | | 5.4% | | | 4.7–6.1 |  | 2.24 | 2.08–2.39 | 0.26 | 6.8% | 6.1–7.5 | .0504 |
| EN | 2.14 | 1.95–2.34 | 0.27 | | 4.2% | | | 3.5–4.8 |  | 2.21 | 2.05–2.38 | 0.25 | 5.8% | 5.1–6.5 | .0072* |
| PN | 1.03 | 0.92–1.14 | 0.06 | | 0.2% | | | 0.1–1.1 |  | 1.16 | 1.06–1.27 | 0.09 | 1.2% | 0.5–2.0 | 1.000 |
| PAF |  |  |  | | 16.0% | | | 14.1–18.0 |  |  |  |  | 24.5% | 22.4–26.6 | < .0072* |
| **PHQ-9** |  |  |  | |  | | |  |  |  |  |  |  |  |  |
| SA | 2.78 | 2.51–3.05 | 0.14 | | 3.3% | | | 2.8–3.7 |  | 2.29 | 2.18–2.42 | 0.14 | 8.0% | 7.4–8.6 | < .0072* |
| PA | 2.48 | 2.31–2.65 | 0.28 | | 7.0% | | | 6.3–7.7 |  | 2.41 | 2.29–2.54 | 0.25 | 6.2% | 5.7–6.7 | 1.000 |
| EA | 3.32 | 3.10–3.54 | 0.26 | | 8.2% | | | 7.6–8.8 |  | 2.93 | 2.80–3.07 | 0.26 | 10.2% | 9.6–10.7 | < .0072* |
| EN | 3.06 | 2.86–3.27 | 0.26 | | 7.7% | | | 7.0–8.3 |  | 2.96 | 2.81–3.11 | 0.25 | 8.9% | 8.3–9.5 | .0504 |
| PN | 1.30 | 1.19–1.41 | 0.07 | | 2.3% | | | 1.5–3.2 |  | 1.39 | 1.30–1.48 | 0.10 | 2.8% | 2.2–3.4 | 1.000 |
| PAF |  |  |  | | 28.4% | | | 26.7–30.2 |  |  |  |  | 36.1% | 34.5–37.7 | < .0072* |

*Note.* RR = Risk Ratio. C = Communality. PAF = Population Attributable Fractions. PD = Physician’s diagnosis of depression. MINI = Mini International Neuropsychiatric Interview. PHQ-9 = Patient Health Questionnaire. 95% confidence intervals are presented. Sum of individual PAF could differ from overall PAF estimates due to rounding*.* P_adj_ = adjusted *p*-values. * *p_adj_* < .05. Adjusted *p*-values exceeding 1 were reported as *p* = 1.000.

### Table S22. *Results of Sensitivity Analysis for Population Attributable Fractions of Depression Cases Attributable to Childhood Maltreatment with Joint-Exposure Approach*

|  | **PAF point estimate** | **95% CIs** | **p_adj_** |
| --- | --- | --- | --- |
| **Physician’s diagnosis of depression** |  |  |  |
| Total  Female  Male | 22.83%  24.01%  19.22% | 21.99%-23.65%  22.91%-25.09%  17.82%-20.58% | <.001** |
| **MINI** |  |  |  |
| Total  Female  Male | 19.17%  21.23%  16.47% | 18.16%-20.99%  19.23%-23.16%  14.33%-18.52% | <.001** |
| **PHQ-9** |  |  |  |
| Total  Female  Male | 29.03%  30.39%  26.08% | 27.83%-30.19%  28.84%-31.96%  24.30%-27.91% | <.001** |

*Note.* PAF = Population Attributable Fractions. MINI = Mini International Neuropsychiatric Interview. PHQ-9 = Patient Health Questionnaire. P_adj =_ adjusted p-values. 95% confidence intervals are presented. p-values refer to sex difference.

### Table S23. *Results of the Sensitivity Analysis of Population Attributable Fractions and Risk Ratios of Depression Cases Attributable to Childhood Maltreatment for the Total Sample*

|  | RR | | | C | PAF | |  | Indiv. weighted PAF | |
| --- | --- | --- | --- | --- | --- | --- | --- | --- | --- |
|  |  | | 95 % CI |  |  | 95 % CI |  |  | 95 % CI |
| **Physician’s diagnosis of depression** |  |  | |  |  |  |  |  |  |
| Sexual Abuse | 2.39 | | 2.31–2.46 | 0.14 | 7.7% | 7.3–8.2 |  | 5.9% | 5.6–6.2 |
| Physical Abuse | 2.01 | | 1.94–2.07 | 0.26 | 7.4% | 7.0–8.0 |  | 4.9% | 4.6–5.2 |
| Emotional Abuse | 2.55 | | 2.48–2.63 | 0.25 | 11.1% | 10.7–11.7 |  | 7.4% | 7.1–7.8 |
| Emotional Neglect | 2.45 | | 2.37–2.52 | 0.26 | 9.6% | 9.1–10.1 |  | 6.4% | 6.1–6.7 |
| Physical Neglect | 1.15 | | 1.10–1.19 | 0.08 | 1.4% | 1.0–1.8 |  | 1.1% | 0.8–1.5 |
| Overall PAF |  | |  |  |  |  |  | 25.8% | 25.0–26.8 |
| **MINI** |  | |  |  |  |  |  |  |  |
| Sexual Abuse | 2.19 | | 2.05–2.33 | 0.13 | 6.0% | 5.3–6.7 |  | 4.7% | 4.2–5.3 |
| Physical Abuse | 1.91 | | 1.80–2.03 | 0.27 | 6.4% | 5.6–7.2 |  | 4.3% | 3.8–4.8 |
| Emotional Abuse | 2.42 | | 2.29–2.56 | 0.26 | 9.4% | 8.6–10.3 |  | 6.4% | 5.9–7.0 |
| Emotional Neglect | 2.20 | | 2.07–2.34 | 0.26 | 7.4% | 6.6–8.2 |  | 5.0% | 4.5–5.5 |
| Physical Neglect | 1.07 | | 1.00–1.16 | 0.08 | 0.7% | 0.1–1.4 |  | 0.6% | 0.1–1.2 |
| Overall PAF |  | |  |  |  |  |  | 21.0% | 19.5–22.5 |
| **PHQ-9** |  | |  |  |  |  |  |  |  |
| Sexual Abuse | 2.55 | | 2.43–2.68 | 0.14 | 8.6% | 7.9–9.2 |  | 6.3% | 5.9–6.8 |
| Physical Abuse | 2.38 | | 2.28–2.49 | 0.26 | 10.0% | 9.3–10.8 |  | 6.4% | 5.9–6.8 |
| Emotional Abuse | 3.11 | | 2.99–3.24 | 0.25 | 14.6% | 13.9–15.4 |  | 9.5% | 9.0–9.9 |
| Emotional Neglect | 2.95 | | 2.82–3.08 | 0.26 | 12.6% | 11.8–13.3 |  | 8.1% | 7.6–8.5 |
| Physical Neglect | 1.28 | | 1.21–1.35 | 0.08 | 2.6% | 2.0–3.3 |  | 2.1% | 1.6–2.6 |
| Overall PAF |  | |  |  |  |  |  | 32.4% | 31.1–33.6 |

*Note.* Sensitivity analysis with relative income position as additional covariate. RR = Risk Ratio. C = Communality. PAF = Population Attributable Fractions. MINI = Mini International Neuropsychiatric Interview. PHQ-9 = Patient Health Questionnaire. 95% confidence intervals are reported. Sum of individual PAF could differ from overall PAF estimates due to rounding*.*

### Table S24. *Results of the Sensitivity Analysis of Population Attributable Fractions and Risk Ratios of Depression Cases Attributable to Childhood Maltreatment for Females*

|  | RR | | | C | PAF | |  | Indiv. weighted PAF | |
| --- | --- | --- | --- | --- | --- | --- | --- | --- | --- |
|  |  | | 95 % CI |  |  | 95 % CI |  |  | 95 % CI |
| **Physician’s diagnosis of depression** |  |  | |  |  |  |  |  |  |
| Sexual Abuse | 2.02 | | 1.94–2.10 | 0.14 | 9.0% | 8.3–9.7 |  | 6.9% | 6.4–7.3 |
| Physical Abuse | 2.06 | | 1.98–2.15 | 0.25 | 7.8% | 7.1–8.3 |  | 5.1% | 4.7–5.5 |
| Emotional Abuse | 2.35 | | 2.26–2.43 | 0.25 | 10.4% | 9.8–10.9 |  | 6.9% | 6.5–7.2 |
| Emotional Neglect | 2.35 | | 2.26–2.44 | 0.25 | 10.2% | 9.5–10.8 |  | 6.7% | 6.4–7.2 |
| Physical Neglect | 1.24 | | 1.19–1.30 | 0.10 | 2.3% | 1.8–2.8 |  | 1.8% | 1.4–2.2 |
| Overall PAF |  | |  |  |  |  |  | 28.5% | 26.2–28.5 |
| **MINI** |  | |  |  |  |  |  |  |  |
| Sexual Abuse | 1.97 | | 1.82–2.12 | 0.13 | 8.1% | 6.9–9.3 |  | 6.4% | 5.5–7.2 |
| Physical Abuse | 1.96 | | 1.81–2.12 | 0.26 | 6.7% | 5.7–7.8 |  | 4.5% | 3.8–5.2 |
| Emotional Abuse | 2.24 | | 2.08–2.40 | 0.26 | 10.4% | 9.2–11.6 |  | 6.9% | 6.1–7.7 |
| Emotional Neglect | 2.18 | | 2.02–2.36 | 0.26 | 8.5% | 7.4–9.7 |  | 5.7% | 5.0–6.5 |
| Physical Neglect | 1.14 | | 1.04–1.25 | 0.09 | 1.3% | 0.3–2.3 |  | 1.1% | 0.3–1.9 |
| Overall PAF |  | |  |  |  |  |  | 24.5% | 22.3–26.8 |
| **PHQ-9** |  | |  |  |  |  |  |  |  |
| Sexual Abuse | 2.28 | | 2.17–2.41 | 0.14 | 11.0% | 10.1–12.0 |  | 8.1% | 7.4–8.8 |
| Physical Abuse | 2.39 | | 2.25–2.53 | 0.25 | 9.8% | 8.9–10.8 |  | 6.2% | 5.7–6.8 |
| Emotional Abuse | 2.89 | | 2.75–3.03 | 0.25 | 16.0% | 15.0–17.0 |  | 10.1% | 9.6–10.7 |
| Emotional Neglect | 2.87 | | 2.72–3.03 | 0.25 | 13.6% | 12.6–14.6 |  | 8.6% | 8.0–9.2 |
| Physical Neglect | 1.33 | | 1.24–1.42 | 0.10 | 3.1% | 2.3–3.9 |  | 2.4% | 1.8–3.0 |
| Overall PAF |  | |  |  |  |  |  | 35.5% | 33.8–37.2 |

*Note.* Sensitivity analysis with relative income position as additional covariate. RR = Risk Ratio. C = Communality. PAF = Population Attributable Fractions. MINI = Mini International Neuropsychiatric Interview. PHQ-9 = Patient Health Questionnaire. 95% confidence intervals are presented. Sum of individual PAF could differ from overall PAF estimates due to rounding*.*

### Table S25. *Results of the Sensitivity Analysis of Population Attributable Fractions and Risk Ratios of Depression Cases Attributable to Childhood Maltreatment for Males*

|  | RR | | | C | PAF | |  | Indiv. weighted PAF | |
| --- | --- | --- | --- | --- | --- | --- | --- | --- | --- |
|  |  | | 95 % CI |  |  | 95 % CI |  |  | 95 % CI |
| **Physician’s diagnosis** |  |  | |  |  |  |  |  |  |
| Sexual Abuse | 2.51 | | 2.32–2.71 | 0.14 | 3.7% | 3.2–4.2 |  | 3.0% | 2.6–3.3 |
| Physical Abuse | 2.01 | | 1.90–2.12 | 0.28 | 7.7% | 6.9–8.5 |  | 5.1% | 4.6–5.6 |
| Emotional Abuse | 2.61 | | 2.46–2.76 | 0.26 | 9.1% | 8.3–9.8 |  | 6.2% | 5.7–6.7 |
| Emotional Neglect | 2.45 | | 2.32–2.59 | 0.26 | 8.5% | 7.8–9.3 |  | 5.8% | 5.3–6.3 |
| Physical Neglect | 1.02 | | 0.95–1.09 | 0.07 | 0.2% | 0.4–0.8 |  | 0.1% | 0.4–0.7 |
| Overall PAF |  | |  |  |  |  |  | 20.9% | 18.9–21.6 |
| **MINI** |  | |  |  |  |  |  |  |  |
| Sexual Abuse | 2.18 | | 1.89–2.50 | 0.12 | 2.5% | 1.8–3.2 |  | 2.1% | 1.5–2.6 |
| Physical Abuse | 1.88 | | 1.71–2.07 | 0.29 | 6.2% | 5.0–7.5 |  | 4.2% | 3.4–5.0 |
| Emotional Abuse | 2.51 | | 2.28–2.74 | 0.27 | 7.8% | 6.7–8.8 |  | 5.4% | 4.6–6.1 |
| Emotional Neglect | 2.10 | | 1.89–2.31 | 0.27 | 5.8% | 4.7–6.9 |  | 4.0% | 3.3–4.7 |
| Physical Neglect | 1.00 | | 0.89–1.12 | 0.06 | 0.1% | 0.0–1.0 |  | 0.1% | 0.0–0.9 |
| Overall PAF |  | |  |  |  |  |  | 16.0% | 13.6–17.6 |
| **PHQ-9** |  | |  |  |  |  |  |  |  |
| Sexual Abuse | 2.67 | | 2.40–2.96 | 0.14 | 4.1% | 3.4–4.8 |  | 3.2% | 2.7–3.7 |
| Physical Abuse | 2.41 | | 2.23–2.60 | 0.28 | 10.5% | 9.2–11.6 |  | 6.7% | 6.0–7.5 |
| Emotional Abuse | 3.25 | | 3.01–3.48 | 0.26 | 12.2% | 11.1–13.3 |  | 8.1% | 7.4–8.7 |
| Emotional Neglect | 2.92 | | 2.71–3.14 | 0.26 | 11.0% | 9.9–12.1 |  | 7.2% | 6.5–7.9 |
| Physical Neglect | 1.21 | | 1.11–1.32 | 0.07 | 2.0% | 1.0–3.0 |  | 1.7% | 0.9–2.5 |
| Overall PAF |  | |  |  |  |  |  | 26.9% | 24.9–28.8 |

*Note.* Sensitivity analysis with relative income position as additional covariate. RR = Risk Ratios. C = Communality. PAF = Population Attributable Fractions. MINI = Mini International Neuropsychiatric Interview. PHQ-9 = Patient Health Questionnaire. 95% confidence intervals are presented. Sum of individual PAF could differ from overall PAF estimates due to rounding*.*

### Table S26. *Sex Differences of the Sensitivity Analysis in Population Attributable Fractions of Depression Cases Attributable to Childhood Maltreatment*

|  | Males | | | | | | | |  | Females | | | | |  |
| --- | --- | --- | --- | --- | --- | --- | --- | --- | --- | --- | --- | --- | --- | --- | --- |
|  | RR | | C | | | indiv. weighted PAF | | |  | RR | | C | indiv. weighted PAF | | *P_adj_* |
|  |  | 95 % CI |  | | |  | | 95 % CI |  |  | 95 % CI |  |  | 95 % CI |  |
| **PD** |  | | |  |  | |  |  |  |  |  |  |  |  |  |
| SA | 2.51 | 2.32–2.71 | 0.14 | | | 3.0% | | 2.6–3.3 |  | 2.02 | 1.94–2.10 | 0.14 | 6.9% | 6.4–7.3 | < .0072* |
| PA | 2.01 | 1.90–2.12 | 0.28 | | | 5.1% | | 4.6–5.6 |  | 2.06 | 1.98–2.15 | 0.25 | 5.1% | 4.7–5.5 | 1.000 |
| EA | 2.61 | 2.46–2.76 | 0.26 | | | 6.2% | | 5.7–6.7 |  | 2.35 | 2.26–2.43 | 0.25 | 6.9% | 6.5–7.2 | < .0072* |
| EN | 2.45 | 2.32–2.59 | 0.26 | | | 5.8% | | 5.3–6.3 |  | 2.35 | 2.26–2.44 | 0.25 | 6.7% | 6.4–7.2 | .0672 |
| PN | 1.02 | 0.95–1.09 | 0.07 | | | 0.1% | | 0.4–0.7 |  | 1.24 | 1.19–1.30 | 0.10 | 1.8% | 1.4–2.2 | < .0072* |
| PAF |  |  |  | | | 20.9% | | 18.9–21.6 |  |  |  |  | 28.5% | 26.2–28.5 | < .0072* |
| **MINI** |  |  |  | | |  | |  |  |  |  |  |  |  |  |
| SA | 2.18 | 1.89–2.50 | 0.12 | | | 2.1% | | 1.5–2.6 |  | 1.97 | 1.82–2.12 | 0.13 | 6.4% | 5.5–7.2 | < .0072* |
| PA | 1.88 | 1.71–2.07 | 0.29 | | | 4.2% | | 3.4–5.0 |  | 1.96 | 1.81–2.12 | 0.26 | 4.5% | 3.8–5.2 | 1.000 |
| EA | 2.51 | 2.28–2.74 | 0.27 | | | 5.4% | | 4.6–6.1 |  | 2.24 | 2.08–2.40 | 0.26 | 6.9% | 6.1–7.7 | .0216* |
| EN | 2.10 | 1.89–2.31 | 0.27 | | | 4.0% | | 3.3–4.7 |  | 2.18 | 2.02–2.36 | 0.26 | 5.7% | 5.0–6.5 | .0072* |
| PN | 1.00 | 0.89–1.12 | 0.06 | | | 0.1% | | 0.0–0.9 |  | 1.14 | 1.04–1.25 | 0.09 | 1.1% | 0.3–1.9 | 1.000 |
| PAF |  |  |  | | | 16.0% | | 13.6–17.6 |  |  |  |  | 24.5% | 22.3–26.8 | < .0072* |
| **PHQ-9** |  |  |  | | |  | |  |  |  |  |  |  |  |  |
| SA | 2.67 | 2.40–2.96 | 0.14 | | | 3.2% | | 2.7–3.7 |  | 2.28 | 2.17–2.41 | 0.14 | 8.1% | 7.4–8.8 | < .0072* |
| PA | 2.41 | 2.23–2.60 | 0.28 | | | 6.7% | | 6.0–7.5 |  | 2.39 | 2.25–2.53 | 0.25 | 6.2% | 5.7–6.8 | 1.000 |
| EA | 3.25 | 3.01–3.48 | 0.26 | | | 8.1% | | 7.4–8.7 |  | 2.89 | 2.75–3.03 | 0.25 | 10.1% | 9.6–10.7 | < .0072* |
| EN | 2.92 | 2.71–3.14 | 0.26 | | | 7.2% | | 6.5–7.9 |  | 2.87 | 2.72–3.03 | 0.25 | 8.6% | 8.0–9.2 | < .0072* |
| PN | 1.21 | 1.11–1.32 | 0.07 | | | 1.7% | | 0.9–2.5 |  | 1.33 | 1.24–1.42 | 0.10 | 2.4% | 1.8–3.0 | 1.000 |
| PAF |  |  |  | | | 26.9% | | 24.9–28.8 |  |  |  |  | 35.5% | 33.8–37.2 | < .0072* |

*Note.* Sensitivity analysis with relative income position as additional covariate. RR = Risk Ratio. C = Communality. PAF = Population Attributable Fractions. PD = Physician’s diagnosis of depression. MINI = Mini International Neuropsychiatric Interview. PHQ-9 = Patient Health Questionnaire. 95% confidence intervals are presented. Sum of individual PAF could differ from overall PAF estimates due to rounding*.* P_adj_ = adjusted *p*-values. * *p_adj_* < .05. Adjusted *p*-values exceeding 1 were reported as *p* = 1.000.

### Table S27. *Sensitivity Analysis of Population Attributable Fractions and Risk Ratios of Depression Cases Attributable to Childhood Maltreatment in L2 Participants Only*

|  | RR | | | C | PAF | |  | Indiv. weighted PAF | |
| --- | --- | --- | --- | --- | --- | --- | --- | --- | --- |
|  |  | | 95 % CI |  |  | 95 % CI |  |  | 95 % CI |
| **Physician’s diagnosis of depression** |  |  | |  |  |  |  |  |  |
| Sexual Abuse | 2.50 | | 2.34–2.67 | 0.14 | 7.4% | 6.6–8.2 |  | 5.8% | 5.2–6.4 |
| Physical Abuse | 2.01 | | 1.89–2.14 | 0.27 | 7.0% | 6.2–7.9 |  | 4.6% | 4.1–5.2 |
| Emotional Abuse | 2.57 | | 2.42–2.73 | 0.26 | 10.2% | 9.4–11.2 |  | 6.9% | 6.3–7.5 |
| Emotional Neglect | 2.49 | | 2.34–2.64 | 0.26 | 9.0% | 8.1–9.9 |  | 6.0% | 5.5–6.6 |
| Physical Neglect | 1.08 | | 1.00–1.17 | 0.07 | 0.7% | 0.1–1.5 |  | 0.6% | 0.1–1.3 |
| Overall PAF |  | |  |  |  |  |  | 23.9% | 22.3–25.6 |
| **PHQ-9** |  | |  |  |  |  |  |  |  |
| Sexual Abuse | 2.65 | | 2.42–2.90 | 0.14 | 8.1% | 7.0–9.2 |  | 6.1% | 5.3–6.9 |
| Physical Abuse | 2.40 | | 2.19–2.61 | 0.27 | 9.4% | 8.1–10.7 |  | 6.0% | 5.2–6.7 |
| Emotional Abuse | 3.24 | | 3.01–3.49 | 0.26 | 14.0% | 12.6–15.4 |  | 9.0% | 8.2–9.8 |
| Emotional Neglect | 3.10 | | 2.85–3.37 | 0.26 | 12.3% | 10.9–13.7 |  | 7.9% | 7.0–8.7 |
| Physical Neglect | 1.38 | | 1.24–1.53 | 0.07 | 3.4% | 2.2–4.7 |  | 2.7% | 1.8–3.7 |
| Overall PAF |  | |  |  |  |  |  | 31.6% | 29.3–34.0 |

*Note.* RR = Risk Ratio. C = Communality. PAF = Population Attributable Fractions. MINI = Mini International Neuropsychiatric Interview. PHQ-9 = Patient Health Questionnaire. 95% confidence intervals are reported. Sum of individual PAF could differ from overall PAF estimates due to rounding*.*

### Table S28. *Sensitivity Analysis of Population Attributable Fractions and Risk Ratios of Depression Cases Attributable to Childhood Maltreatment in L2 Participants Only for Females*

|  | RR | | | C | PAF | |  | Indiv. weighted PAF | |
| --- | --- | --- | --- | --- | --- | --- | --- | --- | --- |
|  |  | | 95 % CI |  |  | 95 % CI |  |  | 95 % CI |
| **Physician’s diagnosis of depression** |  |  | |  |  |  |  |  |  |
| Sexual Abuse | 2.11 | | 1.95–2.27 | 0.13 | 9.0% | 7.8–10.2 |  | 6.9% | 6.0–7.8 |
| Physical Abuse | 2.08 | | 1.92–2.24 | 0.26 | 7.3% | 6.3–8.4 |  | 4.8% | 4.1–5.5 |
| Emotional Abuse | 2.34 | | 2.18–2.52 | 0.26 | 11.0% | 9.8–12.2 |  | 7.2% | 6.5–8.0 |
| Emotional Neglect | 2.43 | | 2.25–2.61 | 0.25 | 10.1% | 8.8–11.3 |  | 6.7% | 5.9–7.4 |
| Physical Neglect | 1.19 | | 1.07–1.30 | 0.08 | 1.7% | 0.7–2.7 |  | 1.4% | 0.7–2.2 |
| Overall PAF |  | |  |  |  |  |  | 27.0% | 24.3–29.3 |
| **PHQ-9** |  | |  |  |  |  |  |  |  |
| Sexual Abuse | 2.31 | | 2.08–2.56 | 0.13 | 10.5% | 8.8–12.3 |  | 7.8% | 6.6–9.0 |
| Physical Abuse | 2.31 | | 2.06–2.58 | 0.26 | 8.8% | 7.2–10.4 |  | 5.6% | 4.6–6.5 |
| Emotional Abuse | 3.01 | | 2.74–3.32 | 0.26 | 15.6% | 13.8–17.6 |  | 9.8% | 8.8–10.9 |
| Emotional Neglect | 2.88 | | 2.59–3.21 | 0.25 | 12.8% | 11.0–14.8 |  | 8.2% | 7.1–9.4 |
| Physical Neglect | 1.44 | | 1.26–1.63 | 0.09 | 4.0% | 2.4–5.6 |  | 3.1% | 1.9–4.3 |
| Overall PAF |  | |  |  |  |  |  | 34.4% | 31.3–37.6 |

*Note.* RR = Risk Ratio. C = Communality. PAF = Population Attributable Fractions. MINI = Mini International Neuropsychiatric Interview. PHQ-9 = Patient Health Questionnaire. 95% confidence intervals are presented. Sum of individual PAF could differ from overall PAF estimates due to rounding*.*

### Table S29. *Sensitivity Analysis of Population Attributable Fractions and Risk Ratios of Depression Cases Attributable to Childhood Maltreatment in L2 Participants Only for Males*

|  | RR | | | C | PAF | |  | Indiv. weighted PAF | |
| --- | --- | --- | --- | --- | --- | --- | --- | --- | --- |
|  |  | | 95 % CI |  |  | 95 % CI |  |  | 95 % CI |
| **Physician’s diagnosis** |  |  | |  |  |  |  |  |  |
| Sexual Abuse | 2.67 | | 2.29–3.07 | 0.12 | 3.5% | 2.7–4.4 |  | 2.9% | 2.2–3.6 |
| Physical Abuse | 2.00 | | 1.80–2.22 | 0.28 | 7.0% | 5.6–8.4 |  | 4.7% | 3.8–5.6 |
| Emotional Abuse | 2.66 | | 2.39–2.93 | 0.27 | 8.5% | 7.2–9.8 |  | 5.8% | 5.0–6.7 |
| Emotional Neglect | 2.37 | | 2.18–2.64 | 0.27 | 7.2% | 5.9–8.5 |  | 4.9% | 4.1–5.8 |
| Physical Neglect | 0.92 | | 0.80–1.05 | 0.06 | 0.7% | 0.2–1.0 |  | 0.6% | 0.2–1.0 |
| Overall PAF |  | |  |  |  |  |  | 17.7% | 15.4–20.1 |
| **PHQ-9** |  | |  |  |  |  |  |  |  |
| Sexual Abuse | 2.84 | | 2.29–3.40 | 0.12 | 3.9% | 2.7–5.0 |  | 3.0% | 2.1–3.9 |
| Physical Abuse | 2.57 | | 2.23–2.95 | 0.28 | 10.6% | 8.4–12.9 |  | 6.7% | 5.4–8.1 |
| Emotional Abuse | 3.33 | | 2.91–3.77 | 0.27 | 11.5% | 9.7–13.4 |  | 7.5% | 6.4–8.7 |
| Emotional Neglect | 3.26 | | 2.83–3.73 | 0.27 | 11.3% | 9.3–13.4 |  | 7.4% | 6.1–8.7 |
| Physical Neglect | 1.30 | | 1.09–1.53 | 0.06 | 2.7% | 0.8–4.6 |  | 2.2% | 0.7–3.8 |
| Overall PAF |  | |  |  |  |  |  | 26.9% | 23.6–30.2 |

*Note.* RR = Risk Ratios. C = Communality. PAF = Population Attributable Fractions. MINI = Mini International Neuropsychiatric Interview. PHQ-9 = Patient Health Questionnaire. 95% confidence intervals are presented. Sum of individual PAF could differ from overall PAF estimates due to rounding*.*

### Table S30. *Disparity Decomposition of* *Sex Differences in Depression by Childhood Maltreatment*

|  | **Physician’s diagnosis** | **MINI classification** | **PHQ-9** |
| --- | --- | --- | --- |
| Female risk | 17.7% | 17.6% | 9.0% |
| Male risk | 10.1% | 11.9% | 5.9% |
| Female-male difference | 7.6% | 5.7% | 3.1% |
| Exposure component | 1.8% | 1.7% | 1.2% |
| Interaction component | -0.3% | -0.4% | -0.5% |
| Total maltreatment-related component | 1.5% | 1.2% | 0.7% |
| Remaining gap | 6.1% | 4.5% | 2.4% |
| % exposure | 23.3% | 29.2% | 37.0% |
| % interaction | -4.2% | -7.4% | -14.6% |
| % accounted for by maltreatment | 19.2% | 21.8% | 22.4% |

*Note.* Female and male risk = standardized predicted depression risks for females and males. Female–male difference = absolute difference in predicted risk between females and males. Exposure component = portion of the disparity attributable to differences in the joint distribution of childhood maltreatment between females and males. Interaction component = contribution of sex differences in the associations between childhood maltreatment and depression (sex × maltreatment interactions). Total maltreatment-related component = sum of the exposure and interaction components. Remaining gap = the portion of the female–male disparity remaining after accounting for childhood maltreatment. Percentage values (% exposure, % interaction, and % explained by maltreatment) are expressed relative to the observed female–male difference in predicted depression risk. Sexual abuse, physical abuse, emotional abuse, physical neglect, and emotional neglect were included as parallel binary exposure variables. Models were adjusted for age, age², and education. Component values and percentages may not sum exactly to the reported totals because of rounding.
