## Supplementary figures and images for "Examining the Contribution of Childhood Maltreatment to the Sex Gap in Depression: Insights from the German National Cohort (NAKO)"

### Supplemental Figure 1

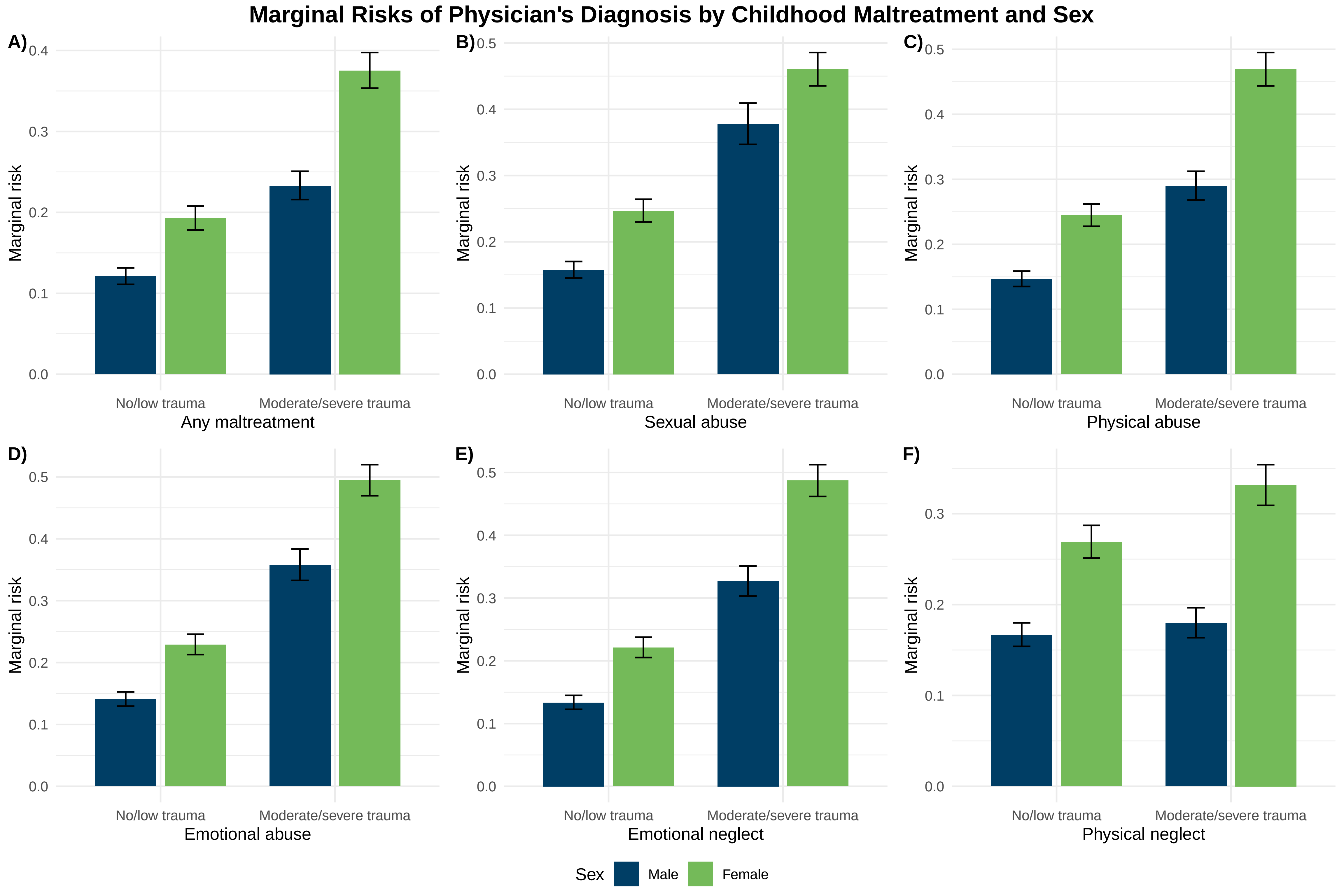

### Supplemental Figure 2

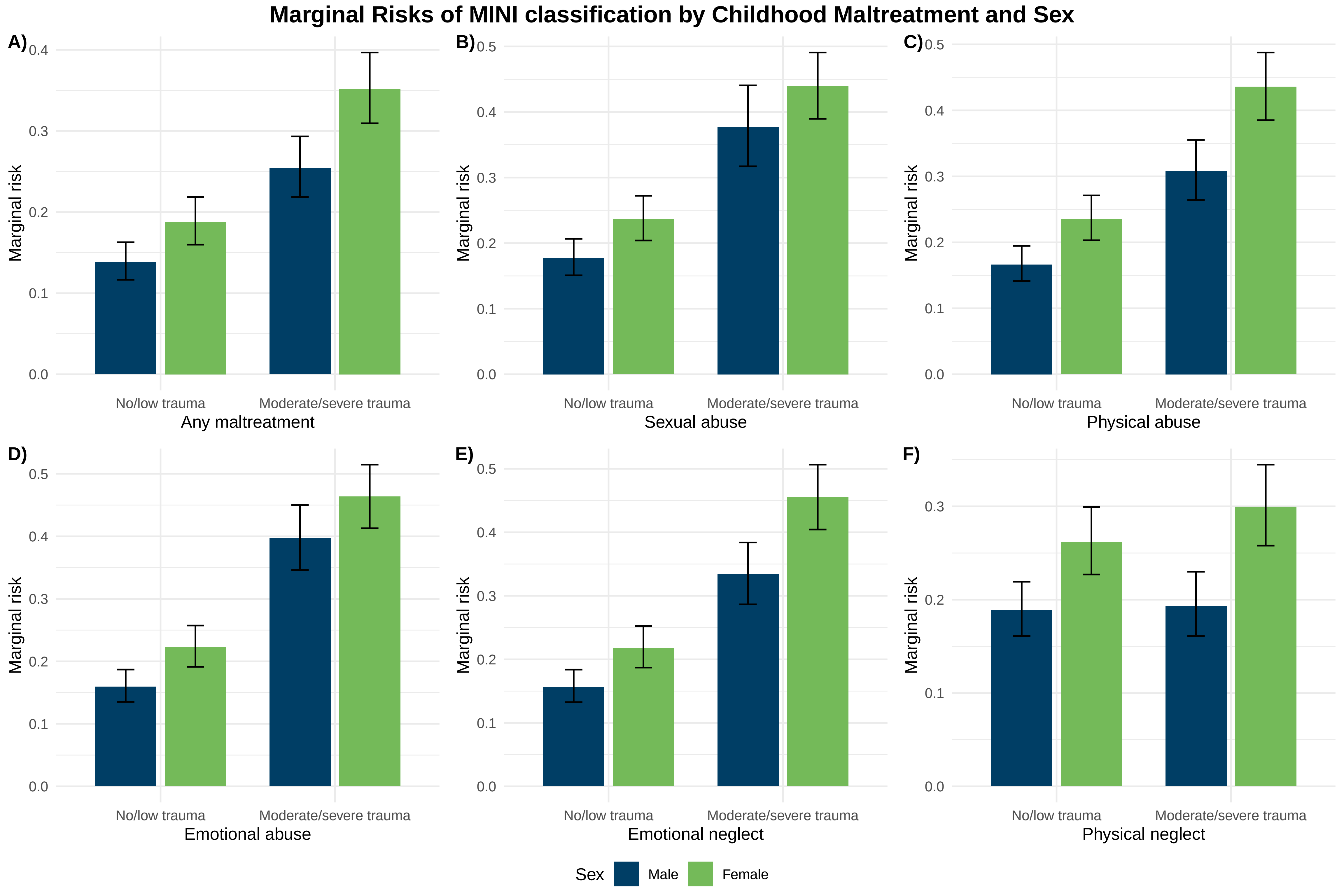

### Supplemental Figure 3

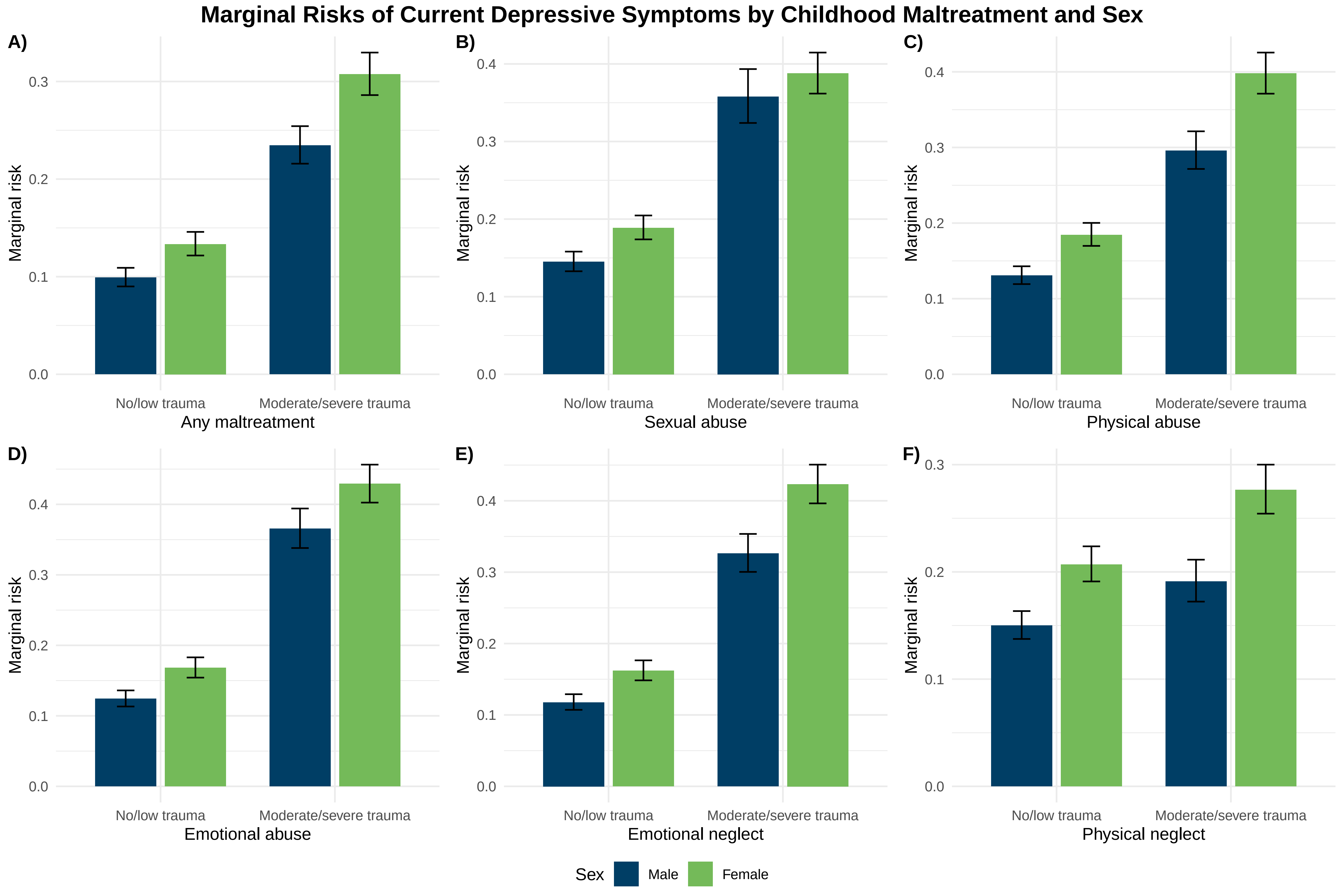
